# Neurobehavioral Signatures in Overgrowth Intellectual Disability Syndromes: Dissecting Genotype-Phenotype Relationships in the PI3K-AKT-MTOR Pathway

**DOI:** 10.1101/2025.08.19.25333388

**Authors:** Aaron D. Besterman, Gerhard Hellemann, Irma Gutierrez-Mejia, Dzung C. Nguyen, Joshua Sadik, Veronica Gandara, Jonathan A. Bernstein, Thomas Frazier, Antonio Y. Hardan, Charis Eng, Mustafa Sahin, Julian A. Martinez-Agosto, the Developmental Synaptopathies Consortium

## Abstract

Overgrowth intellectual disability syndromes (OGIDs) caused by mutations in the PI3K-AKT-MTOR pathway present significant neurobehavioral challenges. While PTEN Hamartoma Tumor Syndrome (PHTS) has been behaviorally characterized, Smith-Kingsmore Syndrome (SKS) has not, limiting our understanding of shared and unique features across OGIDs.

We conducted comprehensive neurobehavioral assessments in 17 individuals with SKS and compared them to previously characterized cohorts with PHTS (n=74), macrocephaly-associated autism (n=33), and healthy controls (n=32). Assessments included standardized measures of motor coordination, adaptive functioning, social interaction, and executive functioning. We performed genotype-phenotype correlation analyses and developed diagnostic classification models using recursive partitioning.

Individuals with SKS showed significant impairments across multiple domains compared to controls. Compared to the PTEN-ASD group, SKS individuals demonstrated particularly severe deficits in motor coordination and adaptive functioning, while executive functioning and behavioral regulation were similarly impaired. Novel clinical features were identified, including immune dysregulation and chronic constipation in SKS, and notably high rates of neonatal teeth (44.7%) in PHTS. Diagnostic classification models incorporating both behavioral and medical features achieved above-chance accuracy in distinguishing between conditions, with neonatal teeth emerging as a key distinguishing feature for PHTS. Domain-specific analyses showed variants in the PTEN phosphatase domain were associated with more severe social and executive function deficits compared to C2 domain variants. Correlation analyses between variant pathogenicity scores and clinical measures revealed limited consistent associations, though Combined Annotation Dependent Depletion (CADD) scores showed stable correlations with sensory processing measures across cohorts.

Our findings establish distinct neurobehavioral profiles between SKS and PHTS, suggesting different impacts of MTOR versus PTEN mutations on neural circuit development. The identification of novel phenotypic features expands the clinical spectrum of these disorders and provides new diagnostic markers. The limited predictive value of variant pathogenicity scores for neurobehavioral outcomes emphasizes the need for comprehensive individual assessments. These results provide a foundation for developing targeted interventions while highlighting the complexity of genotype-phenotype relationships in PI3K-AKT-MTOR pathway disorders.

## INTRODUCTION

Overgrowth intellectual disability syndromes (OGIDs) represent a diverse group of rare genetic disorders characterized by excessive somatic and cellular growth accompanied by developmental delays, intellectual disability, and other complex medical and neurobehavioral challenges.^1,2^ Many OGIDs result from disruption in genes that are part of the PI3K/AKT/MTOR signaling pathway, a central regulator of cellular growth, metabolism and homeostasis.^3^ Smith-Kingsmore Syndrome (SKS) is an OGID that was first described in 2013^4^ that specifically occurs from activating variants in the *MTOR* gene, leading to somatic overgrowth^5,6^. The core features of SKS include macrocephaly/megalencephaly, intellectual disability, and seizures with a range of other prevalent medical and neurobehavioral comorbidities.^5^ As more individuals with SKS are identified, it will be critical to refine key clinical features of the disorder to establish a natural history. Additionally, SKS families have indicated that neurobehavioral and mental health issues are a top concern of theirs, and it is therefore critical that the neurobehavioral profile of individuals with SKS be delineated.^7^

The potential benefits of such efforts have been demonstrated in a related PI3K/AKT/MTOR OGID, *PTEN* Hamartoma Tumor Syndrome (PHTS). ^8–11^ It was found that individuals with PHTS have reduced performance on measures of attention, impulsivity, reaction time, processing speed, and motor coordination, but in those with co-occurring autism spectrum disorder (ASD) the deficits were more pronounced.^8^ They have subsequently been shown to have relatively stable symptomatology over time and these measures may be used in future research to examine patient outcomes at the individual level as well as to detect negative deviations from the expected trajectory that can be used to inform intervention strategies. ^9^ Unlike PHTS, the neurobehavioral profile of SKS remains unknown. It may be that individuals with SKS will have a similar profile to those with PHTS, as individual with variants in genes that lie within the same signaling pathway (e.g. CHD8, RAS-opathies, GATOR-opathies,) can have meaningful phenotypic and clinical similarities that allow for comparative investigations of gene-to-disease progression. ^12–14^ To further define the clinical and neurobehavioral profile of SKS relative to other OGIDs, we completed a full assessment of 17 individuals with SKS and refine previously reported clinical features of both SKS and PHTS and describe a comprehensive neurobehavioral profile of individuals with SKS for the first time. We directly compare the neurobehavioral profile to four groups 1) Individuals with PTEN variants and ASD (PTEN-ASD), 2) Individuals with PTEN but without ASD (PTEN no-ASD), 3) Individuals with macrocephaly and ASD, but no known genetic etiology (Macro-ASD) and 4) An unaffected control group (Healthy Control). We then performed genotype-phenotype correlation analyses across OGIDs to identify potential relationships between specific genetic variants and neurobehavioral outcomes that could inform patient care and prognosis.

Our comprehensive analysis reveals both shared and distinct neurobehavioral features between SKS and PHTS, highlighting the complex relationship between PI3K/AKT/MTOR pathway disruption and clinical manifestations. By systematically comparing these conditions, we aim to advance understanding of pathway-specific effects on neurodevelopment and establish a foundation for more targeted therapeutic approaches for OGIDs. These insights may also help inform clinical care by identifying which features are likely to be shared across PI3K/AKT/MTOR-related disorders versus those that may be unique to specific genetic etiologies.

## MATERIALS AND METHODS

### SKS Participant Recruitment and Assessment

All SKS participants were recruited through The University of California Los Angeles (UCLA) Medical Center, Los Angeles, United States as part of an institutional review board-approved research study (#12-000989). Informed consent was obtained from all legal guardians of participants due to the participants being minors and/or lacking the capacity to consent to a research study due to intellectual deficits. Participants were identified through advertisement on the SKS Foundation Website (www.smithkingsmore.org), GeneMatcher^15^, from colleague referral, and by word-of-mouth through SKS parent networks. Participants had to have a documented *de novo* variant in *MTOR* reported by a CLIA-certified diagnostic laboratory that was classified as either a variant of unknown significance, likely pathogenic, or pathogenic. Additionally, participants had to have clinical symptoms of overgrowth, most commonly macrocephaly, and atypical neurodevelopment. Participants were given the option of completing a full in-person medical and neurobehavioral evaluation at UCLA or having a packet of self-assessments mailed to them, which they would return with medical records, photos, and genetic test results. Five out of 17 participants completed a full in-person evaluation, and 12 participants completed the at-home assessment battery. Only the assessments that all patients were able to complete through both recruitment approaches were used in the analyses for this study.

The neurobehavioral battery (Table S1) was adapted from the PTEN-DSC battery^8^ to allow for a direct comparison between the two cohorts. All in-person medical assessments were completed by J.A.M.A., a board-certified pediatrician and medical geneticist. All in-person neurobehavioral assessments were completed and scored by experienced and licensed neuropsychologists or psychometrists at the UCLA Center for Autism Research & Treatment. Self-assessments returned by mail were reviewed, scored, and entered into our secured database by I.G.M. All measures were scored according to published test manuals using age- and/or sex-corrected norms as available/appropriate. Medical information was extracted, reviewed, and tabulated by I.G.M., J.S., and A.D.B. Demographic characteristics, including age, sex, race and ethnicity, in comparison to previously reported PHTS cohorts^8^, were collected (Table S2).

### PTEN Patient Recruitment and Assessment

Participants were recruited from four large tertiary medical centers (Cleveland Clinic, Boston Children’s Hospital, Stanford University Medical Center, and University of California, Los Angeles) as part of an institutional review board-approved, ongoing, multicenter prospective study designed to examine the natural history of autism and germline heterozygous PTEN mutations (clinicaltrials.gov: NCT02461446). Details of recruitment, neurobehavioral assessment, and genotyping have been published elsewhere.^8^

### Neurobehavioral Phenotype Comparison

#### Statistical Comparison

Neurobehavioral features were assessed using standardized parent-report measures across seven domains: autism symptoms (SRS-2, RBS-R), adaptive function (VABS-II), behavior (CBCL), motor skills (DCDQ), aberrant behavior (ABC), executive function (BRIEF), and sensory processing (SSP) (Table S1). For measures with multiple versions (BRIEF-P and BRIEF-PK), scores were combined into unified variables. All analyses were performed using R version 4.3. Summary statistics for each measure were calculated as median and interquartile range (IQR) across five clinical groups: SKS, PTEN-ASD, Macrocephaly ASD, PTEN no-ASD, and Healthy Controls (Tables S2 and S3). To handle missing data, calculations were performed using all available data points for each measure. Group comparisons were conducted using three approaches. First, Mann-Whitney U tests were used to compare SKS to healthy controls, providing a baseline assessment of neurobehavioral differences (Table S4). Second, Mann-Whitney U tests compared SKS to PTEN-ASD to specifically examine differences between these two genetic conditions (Table S4). For all pairwise comparisons, effect sizes were calculated as r-values (Z/√N), and p-values were adjusted for multiple comparisons using the false discovery rate (FDR) method within each domain (Table S4). Third, MANOVA was performed to compare features across SKS, PTEN-ASD, and macrocephaly-associated ASD groups, evaluating potential differences among genetic and phenotypic forms of macrocephaly-associated neurodevelopmental conditions (Table S5). For MANOVA analyses, Pillai’s trace was used as the test statistic due to its robustness to violations of assumptions.

#### PCA Analysis

To evaluate whether distinct patterns of neurobehavioral features could differentiate between diagnostic groups, we performed principal component analysis (PCA) on 34 behavioral measures spanning multiple domains: social function (Social Responsiveness Scale-2), repetitive behaviors (Repetitive Behavior Scale-Revised), behavioral problems (Child Behavior Checklist, Aberrant Behavior Checklist), motor skills (Developmental Coordination Disorder Questionnaire), sensory processing (Short Sensory Profile subdomains), executive function (BRIEF-P and BRIEF-PK), and adaptive function (Vineland Adaptive Behavior Scales-2) (Fig 1A). Missing data, which ranged from 19.2% (Short Sensory Profile measures) to 57.7% (Vineland Adaptive Behavior Scales-2 Motor Skills), were handled using mean imputation for each variable. All variables were standardized to zero mean and unit variance before performing PCA. BRIEF-P and BRIEF-PK measures were combined to include only overlapping subdomains (e.g., Global Executive Composite, Inhibition, Shift, Emotional Control, Working Memory, and Plan/Organize). The analysis included 156 subjects across five groups: SKS (n = 17), PTEN-ASD (n = 44), PTEN no-ASD (n = 30), Macrocephaly ASD (n = 33), and Healthy Controls (n = 32). Results are presented as scatter plots of the first two principal components with 95% confidence ellipses for each group (Fig 1A). All analyses were performed using R (version 4.3) with the ‘tidyverse’ package suite. PCA plots were generated using ggplot2 and factoextra packages. Missing or non-numeric entries were carefully examined and excluded where appropriate.

**Figure 1.**
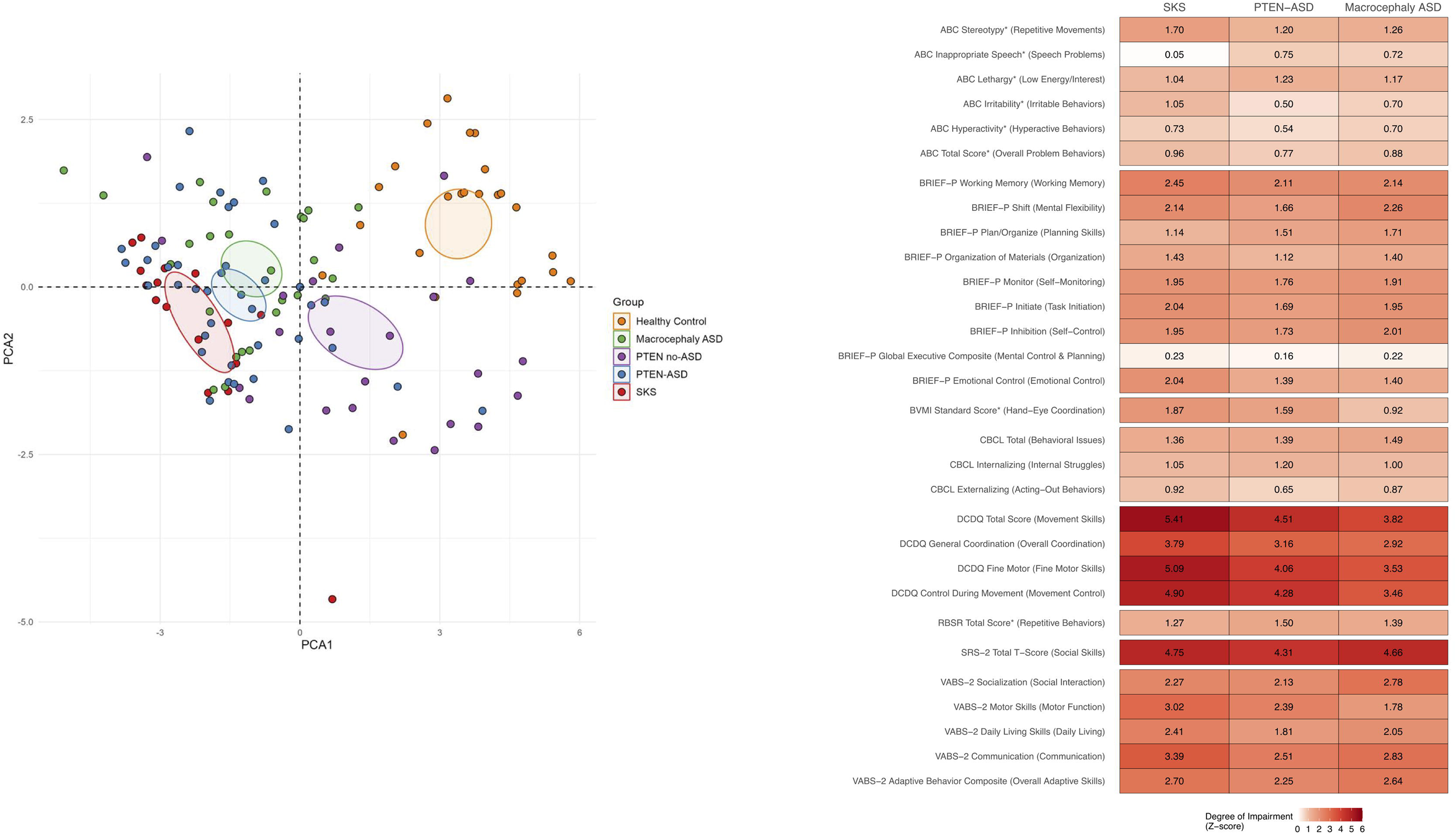
Neurobehavioral Profiles in Patients with OGIDs. (A) Principal component analysis (PCA) of 32 behavioral measures across cognitive, social, motor, and adaptive domains in 140 subjects. Data points represent individual subjects colored by group: SKS (MTOR mutations, red), PTEN-ASD (blue), PTEN no-ASD (green), Macrocephaly ASD (purple), and Healthy Controls (orange). Ellipses indicate 95% confidence regions for each group. PCA was performed on standardized scores with mean imputation for missing values. (B) Heatmap showing the severity of impairment across behavioral domains for SKS, PTEN-ASD, and Macrocephaly ASD groups. Colors indicate z-score magnitude, with darker red representing greater impairment relative to reference groups. Z-scores were calculated using Healthy Controls as reference for BRIEF-P, CBCL, DCDQ, SRS-2, and VABS-2 measures. For measures marked with an asterisk (*), PTEN no-ASD served as the reference group because healthy control data was missing. All scores were oriented so that higher values (darker red) indicate greater impairment. Abbreviations: ABC, Aberrant Behavior Checklist; BRIEF-P, Behavior Rating Inventory of Executive Function-Preschool; BVMI, Beery-Buktenica Visual-Motor Integration; CBCL, Child Behavior Checklist; DCDQ, Developmental Coordination Disorder Questionnaire; RBS-R, Repetitive Behavior Scale-Revised; SRS-2, Social Responsiveness Scale-2; VABS-2, Vineland Adaptive Behavior Scales-2.

#### Heatmap of Z-scores

Z-scores in reference to healthy controls were calculated for all neurobehavioral measures to enable standardized comparison across different assessment instruments. For measures where healthy control data was available (BRIEF, CBCL, DCDQ, SRS-2, SSP, and VABS-2), z-scores were calculated using healthy controls as the reference group. Measures that lacked healthy control data (ABC and RBS-R) were removed from the analysis to ensure consistency in referencing across all measures. For each measure, z-scores were computed as the difference between each individual’s score and the reference group mean, divided by the reference group standard deviation. For measures where higher scores indicate better function (VABS-2, DCDQ, and SSP), z-scores were multiplied by −1 to maintain consistent directionality across all measures, ensuring that higher z-scores represent greater impairment. This standardization facilitates direct comparison across different instruments, with positive z-scores representing greater impairment relative to the reference group. Results were visualized using a heatmap where color intensity represents the magnitude of impairment (degree of deviation from the reference group), with darker red indicating greater impairment. The heatmap displays mean z-scores for three groups: individuals with SKS (MTOR mutations), individuals with PTEN mutations and ASD (PTEN-ASD), and individuals with macrocephaly and ASD (Macrocephaly ASD). All analyses were performed using R (version 4.3) with the ‘tidyverse’ package suite.

### Phenotype-Protein Domain Correlation

We analyzed neurobehavioral phenotypes across MTOR and PTEN protein domains using R version 4.3 with the tidyverse suite of packages. Participants were grouped based on genetic variant location: MTOR FAT domain, MTOR Kinase domain, PTEN C2 domain, or PTEN phosphatase domain. PTEN variant carriers were further stratified by ASD status (PTEN-ASD vs PTEN no-ASD). We assessed differences between groups across multiple standardized neurobehavioral measures, including the DCDQ, SRS-2, VABS-2, ABC, RBS-R, CBCL, and BRIEF. For each measure, we performed a Kruskal-Wallis test to identify significant differences between domain groups (Table S3) followed by pairwise comparisons between domains within each protein (Table S4). The top ten phenotypes showing significant differences (p < 0.05) on the Kruskal-Wallis test were visualized using boxplots with individual data points to show the distribution of scores across protein domains (Fig 2).

**Figure 2.**
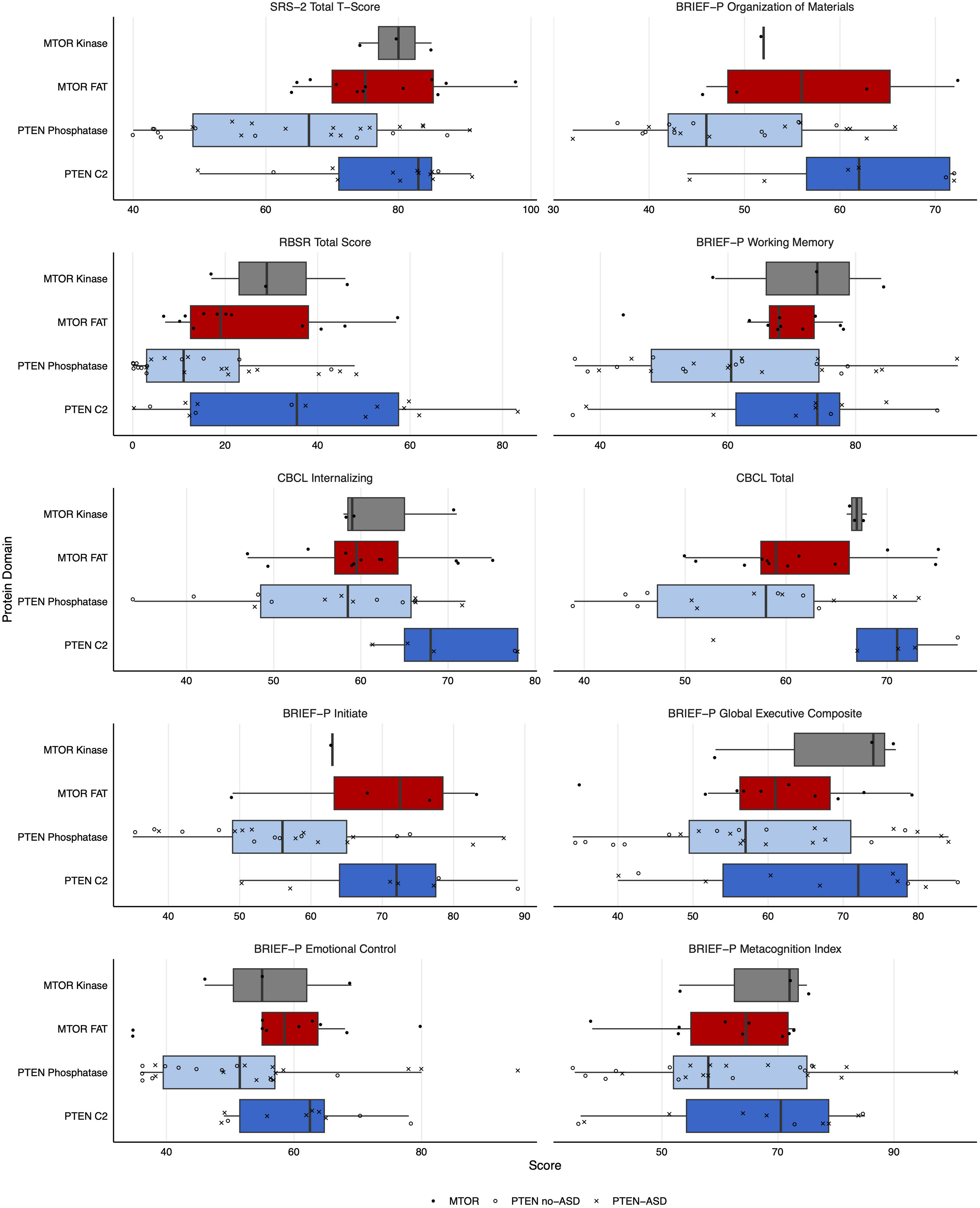
Neurobehavioral Scores Across PTEN and MTOR Domains. Domain-specific differences in top neuropsychological phenotypes. Box plots showing the distribution of scores for the ten most significantly different neuropsychological measures across PTEN and MTOR protein domains. Box plots represent median and interquartile range, with individual data points overlaid. For PTEN variants, filled circles indicate individuals with ASD diagnosis while X’s indicate those without ASD. Diamond shapes represent individuals with MTOR variants. Colors indicate protein domains: PTEN Phosphatase (light blue), PTEN C2 (dark blue), MTOR FAT (light red), and MTOR PI3K (dark red). Abbreviations: SRS, Social Responsiveness Scale; BRIEF-P, Behavior Rating Inventory of Executive Function - Preschool Version; CBCL, Child Behavior Checklist.

### Pathogenicity-Phenotype Correlation

Neurobehavioral phenotype measures were correlated with four different variant pathogenicity scores: RAS^16^, CADD^17^, REVEL^18^, and AlphaMissense^19^. The analysis was conducted across four distinct groups: SKS, PHTS, SFARI genes in the PI3K-AKT-MTOR cluster, and then these three cohorts combined (“Combined OGIDs”). Only missense variants were included in this analysis and all LOF and early termination variants were excluded. For each group, we calculated Pearson correlation coefficients between each neurobehavioral measure and pathogenicity scores. Statistical significance was assessed using a two-tailed test (Table 4).

We conducted correlation analyses between pathogenicity prediction scores (AlphaMissense, CADD, RAS, and REVEL) and neurobehavioral measures across multiple assessment tools (BRIEF, CBCL, DCDQ, SRS-2, SSP, and VABS-2). First, to assess generalizability of findings across multiple OGIDs, we added patient data from SFARI Searchlight which had matching VABS-2, CBCL and SRS-2 phenotype data. A functional network of 159 proteins coded by the SFARI Searchlight v10.0 genes (which included *PTEN*) plus MTOR was created using the ‘multiple proteins’ function in the STRING database (https://string-db.org/)^20^. The clusters were created using the built-in Markov Cluster Algorithm with inflation factor of 2.1. We then extracted the cluster that included *MTOR* and *PTEN* along with the following Simons Searchlight genes: *CTNNB1* (N = 97), *HIVEP2* (N = 65), *PPP2R1A* (N = 16), *PP2R5D* (N = 231), and *WAC* (N = 6) and confirmed enrichment for the PI3K-AKT-MTOR signaling pathway via the reactome knowledgebase (http://reactome.org).^21^ Correlations were analyzed separately for four groups: SKS (MTOR variants), PTEN variants, SFARI PI3K-AKT-MTOR pathway variants (CTNNB1, HIVEP2, PPP2R1A, PP2R5D, and WAC), and a combined cohort of all OGID variants.

Pearson correlation coefficients were calculated for each pathogenicity score-behavioral measure pair. P-values were adjusted for multiple comparisons using the False Discovery Rate (FDR) method (Table 4). To assess the stability of the correlations, we performed bootstrap analysis with 1,000 resamples for each correlation. For each bootstrap iteration, we recorded the correlation coefficient and significance status (p < 0.05). Correlations were classified as “Very Stable” (≥80% significant resamples), “Moderately Stable” (60-79% significant resamples), “Somewhat Stable” (40-59% significant resamples), or “Unstable” (<40% significant resamples) (Fig 3; Supplement 2).

**Figure 3.**
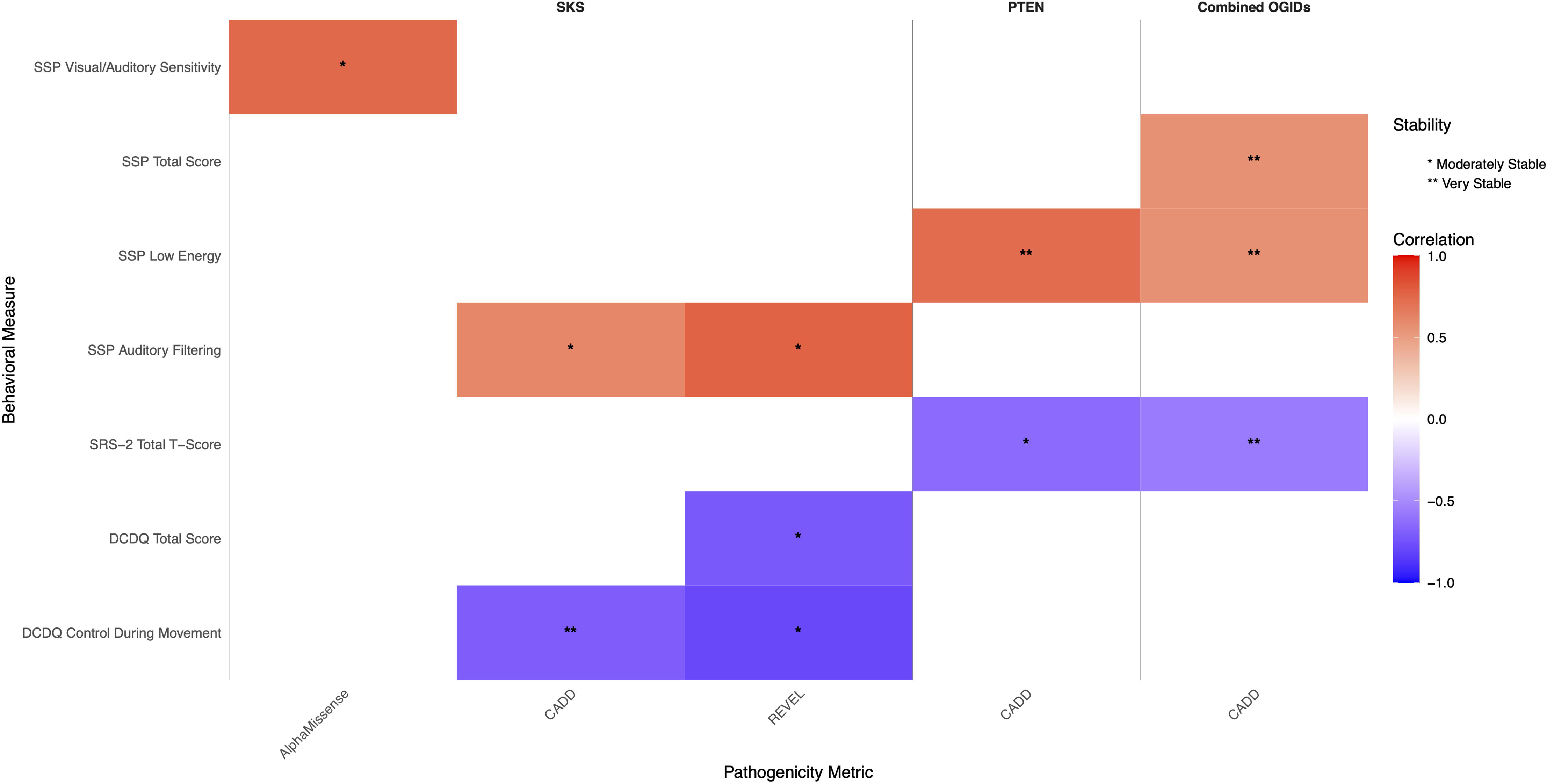
Stable Correlations Between Pathogenicity Metrics and Behavioral Measures in OGIDs. Heatmap displaying only the statistically stable correlations identified through bootstrap analysis (1000 iterations) from the complete correlation analysis shown in Table 4. Colors represent correlation coefficients ranging from −1.0 (dark blue) to +1.0 (dark red), with white indicating no correlation. Asterisks indicate the level of stability (* Moderately Stable, ** Very Stable). The analysis includes three cohorts: SKS, PTEN, and Combined OGIDs (pooled analysis). Behavioral measures shown demonstrated stable correlations with at least one pathogenicity metric (CADD, REVEL, or AlphaMissense) in at least one cohort. Measures without stable correlations are not displayed in the heatmap. Abbreviations: CBCL, Child Behavior Checklist; DCDQ, Developmental Coordination Disorder Questionnaire; SRS-2, Social Responsiveness Scale-2; SSP, Short Sensory Profile; VABS-2, Vineland Adaptive Behavior Scales-2.

### Classification Analysis via Recursive Partitioning

To determine if there were multivariate predictive patterns, we used a machine learning approach – recursive partitioning – to develop decision trees that differentiated between the different cohorts in this sample. We chose recursive partitioning over random forest approaches as this approach generates a single decision tree that can be directly interpreted. While random forest approaches provide potentially marginally higher explanatory power, we felt that the trade off in loss of interpretability was not worth it. To determine the generalizability of model fit we use a 10-fold cross-validated error. Using that approach the data is split into 10 subsamples and the fit of the model is evaluated for each of the subsamples by generating the model on the other 9 subsamples and evaluating its fit on the last subsample. This provides an estimate of how well the model performs on new data, and how much variability we can expect for the model fit across different samples. The raw relative error underestimates the misclassification rate because it is the fit on the same data that the model was developed on.

## RESULTS

### Clinical Phenotypes

#### SKS

Analysis of our SKS cohort (n=17) revealed consistent neurological and developmental manifestations as core features of the syndrome (Table 1). Beyond the universal presence of macrocephaly and developmental delay, we found high rates of hypotonia, dysmorphic features, and seizures. Central nervous system abnormalities were frequent, with nearly half of patients showing ventriculomegaly on neuroimaging. Gastrointestinal symptoms, particularly chronic constipation, emerged as a prominent finding. Sleep disturbance affected a substantial portion of the cohort. We observed a notable pattern of immune system involvement, characterized by recurrent infections and increased prevalence of asthma. Ophthalmologic abnormalities were diverse but consistently present across the cohort.

**Table 1.**
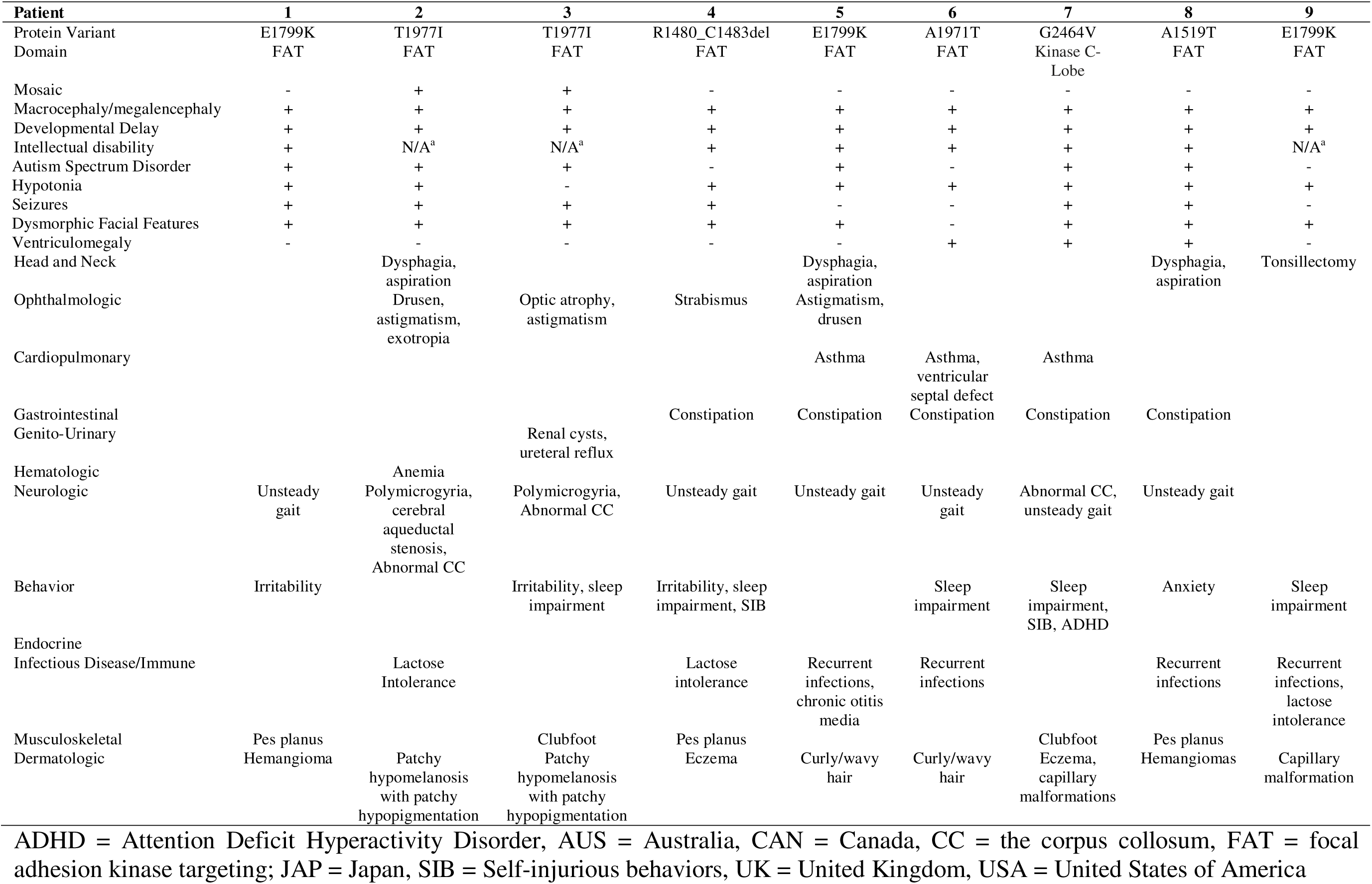
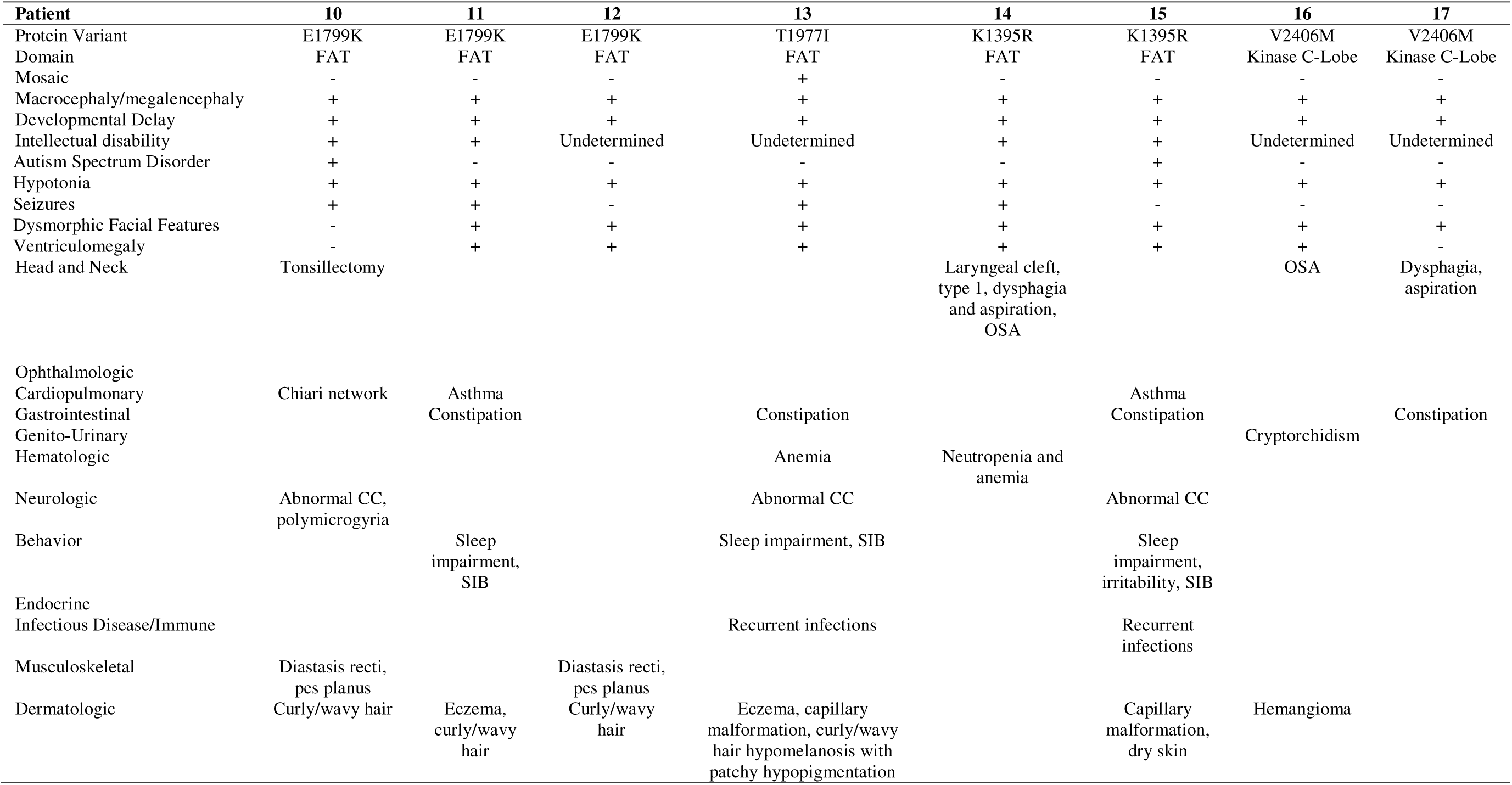
SKS Patient Genotype and Clinical Phenotypes.

**Table 2.** Summary of Published Clinical Phenotypes in Smith Kingsmore Syndrome.

| **Neuropsychiatric** | **N** | **%** |
| --- | --- | --- |
| Macrocephaly | 63/63 | 100 |
| Intellectual disability | 53/55 | 96.4 |
| Central nervous system anomalies (on neuroimaging) | 33/58 | 56.9 |
| Seizures | 34/63 | 54.0 |
| Ventriculomegaly | 28/58 | 48.3 |
| Hypotonia | 29/63 | 46.0 |
| Autism spectrum disorder | 26/63 | 41.3 |
| Speech anomaly | 9/27 | 33.3 |
| Behavioral problems | 19/63 | 30.2 |
| Sleep disturbance | 13/63 | 20.6 |
| Abnormal corpus callosum | 11/63 | 17.5 |
| Neurocognitive deterioration | 9/63 | 14.3 |
| Abnormal gait | 6/63 | 9.5 |
| Polymicrogyria | 5/63 | 7.2 |
| Obsessive behaviors | 2/63 | 3.2 |
| **Ophthalmologic** |  |  |
| Astigmatism | 3/63 | 4.8 |
| Myopia | 2/63 | 3.2 |
| Cerebral visual impairment | 2/63 | 3.2 |
| Iris colobomata | 2/63 | 3.2 |
| Drusen | 2/63 | 3.2 |
| Retinal dysfunction | 1/63 | 1.6 |
| Exotropia | 1/63 | 1.6 |
| Optic atrophy | 1/63 | 1.6 |
| Cataract (adult) | 1/63 | 1.6 |
| **Musculoskeletal** |  |  |
| Macrosomia | 12/27 | 44.4 |
| Pes planus/talipes | 10/63 | 15.9 |
| Diastasis recti | 6/63 | 9.5 |
| Scoliosis | 5/63 | 7.9 |
| Small thorax | 4/63 | 6.3 |
| Joint hypermobility | 4/63 | 6.3 |
| Ulnar deviation of the hands | 1/63 | 1.6 |
| Proximally placed thumbs | 1/63 | 1.6 |
| Bifid distal phalanx of left thumb | 1/63 | 1.6 |
| **Head and Neck** |  |  |
| Obstructive sleep apnea | 3/63 | 4.8 |
| Laryngeal cleft | 1/63 | 1.6 |

| **Endocrine/Immune/Allergy** | **N** | **%** |
| --- | --- | --- |
| Recurrent infections | 9/63 | 14.3 |
| Lactose intolerance | 6/63 | 9.5 |
| Neonatal hypoglycemia | 4/63 | 6.3 |
| Persistent food allergies | 3/63 | 4.8 |
| Hypoglycemia | 2/63 | 3.2 |
| Autoimmune hypothyroidism | 2/63 | 3.2 |
| Hypoglycemia | 2/63 | 3.2 |
| Precocious puberty | 1/63 | 1.6 |
| Insulin dependent diabetes mellitus | 1/63 | 1.6 |
| IgA deficiency | 1/63 | 1.6 |
| **Dermatologic** |  |  |
| Curly/wavy hair | 14/33 | 42.4 |
| Patchy hypomelanosis | 11/63 | 17.5 |
| Capillary malformation of the skin | 10/63 | 15.9 |
| Hemangiomas | 7/63 | 11.1 |
| Café-au-lait spots | 3/63 | 4.8 |
| Angiokeratoma | 1/63 | 1.6 |
| **Gastrointestinal** |  |  |
| Constipation | 11/63 | 17.5 |
| Dysphagia | 7/63 | 11.1 |
| Diarrhea | 3/63 | 4.8 |
| Prominent abdomen | 2/63 | 3.4 |
| Intestinal polyps | 1/63 | 1.6 |
| **Genitourinary** |  |  |
| Cryptorchidism | 3/63 | 4.7 |
| Renal cysts | 2/63 | 3.4 |
| Hypospadias | 1/63 | 1.8 |
| Ureteral reflux | 1/63 | 1.6 |
| **Cardiopulmonary** |  |  |
| Asthma | 9/63 | 14.3 |
| Ventricular septal defect | 1/63 | 1.6 |
| Chiari network | 1/63 | 1.6 |
| **Hematology-Oncology** |  |  |
| Anemia | 5/63 | 7.9 |
| Neutropenia | 1/63 | 1.6 |
| Intraosseous meningiomas (adult) | 1/63 | 1.6 |

#### PHTS

Neurological and developmental features emerged as primary characteristics, with macrocephaly present universally and a markedly high prevalence of autism spectrum disorder (Table 3). The cohort showed substantial early developmental challenges, with nearly a third requiring NICU admission and many experiencing early feeding difficulties. Early childhood manifestations were particularly notable, including a high frequency of neonatal teeth and recurrent otitis media. The phenotype included significant dermatological involvement, primarily manifesting as birthmarks and eczema. Respiratory symptoms were common, particularly asthma and abnormal breathing patterns. Gastrointestinal symptoms, though less frequent than other manifestations, consistently presented as chronic diarrhea in affected individuals. The cohort also demonstrated various musculoskeletal features, with lipomas being the most frequent finding. Notably, many of these clinical features appeared to manifest early in development, suggesting an early-onset, multi-system disorder.

**Table 3.** Summary of Clinical Phenotypes in PTHS from DSC.

| **Neuropsychiatric** | **N** | **%** |
| --- | --- | --- |
| Macrocephaly | 79/79 | 100 |
| Autism Spectrum Disorder | 66/92 | 71.7 |
| Abnormal Gait | 17/90 | 18.9 |
| Seizures | 17/94 | 18.1 |
| Epilepsy | 7/85 | 8.2 |
| Head Trauma | 6/94 | 6.4 |
| Febrile Seizures | 5/92 | 5.4 |
| Meningitis | 2/94 | 2.1 |
| Encephalitis | 2/94 | 2.1 |
| Cerebral Palsy | 1/94 | 1.1 |
| **Symptoms in Infancy** |  |  |
| Hypotonia | 47/92 | 51.1 |
| Extreme Passivity | 30/93 | 32.3 |
| Decreased attention to people | 27/93 | 29.0 |
| Irritation | 26/94 | 27.7 |
| Poor Feeding | 25/92 | 27.2 |
| Disrupted sleep pattern | 24/93 | 25.8 |
| Lethargy | 21/93 | 22.6 |
| Decreased Alertness | 20/92 | 21.7 |
| **Musculoskeletal** |  |  |
| Lipoma | 12/94 | 12.8 |
| Spinal Abnormalities | 8/94 | 8.5 |
| **Dermatologic** |  |  |
| Birthmarks | 44/94 | 46.8 |
| Eczema | 27/92 | 29.3 |
| Skin Infections | 11/94 | 11.7 |
| Penile freckling | 4/94 | 4.3 |
| Hyperpigmentation | 3/94 | 3.2 |
| Capillary malformation | 2/94 | 2.1 |
| Hypopigmentation | 1/94 | 1.1 |
| **Head and Neck** |  |  |
| Neonatal teeth | 42/94 | 44.7 |
| Recurrent otitis media | 38/94 | 40.4 |
| Ear tubes | 29/94 | 30.9 |
| Dental abnormalities | 17/94 | 18.1 |
| Strabismus | 11/94 | 11.7 |
| Deafness | 4/93 | 4.3 |
| Cleft palate | 2/94 | 2.1 |
| **Gastrointestinal** |  |  |
| Diarrhea | 25/94 | 26.6 |
| **Cardiopulmonary** |  |  |
| Asthma | 24/94 | 25.5 |
| Abnormal breathing | 23/94 | 24.5 |
| Cardiac defect | 12/93 | 12.9 |
| Aspiration | 5/94 | 5.3 |
| Lung malformation | 2/93 | 2.2 |

| **Neonatal Complications** | **N** | **%** |
| --- | --- | --- |
| Neonatal intensive care unit | 29/94 | 30.9 |
| Jaundice | 18/94 | 19.1 |
| Cyanosis | 8/94 | 8.5 |
| Hypoglycemia | 5/94 | 5.3 |
| Meconium aspiration | 2/94 | 2.1 |
| **Pregnancy Complications** |  |  |
| Preeclampsia | 8/94 | 8.5 |
| Gestational diabetes | 7/94 | 7.4 |
| Polyhydramnios | 2/94 | 2.1 |
| Placental abruption | 2/94 | 2.1 |
| Cervical funneling | 2/94 | 2.1 |
| Vasa previa | 1/94 | 1.1 |
| **Diet** |  |  |
| Food allergy | 15/94 | 16.0 |
| Food texture aversion | 12/94 | 12.8 |
| Gluten free | 12/94 | 12.8 |
| Dairy free | 10/94 | 10.6 |
| Food temperature aversion | 5/94 | 5.3 |
| Blended Food | 5/94 | 5.3 |
| Fine chopped food | 5/94 | 5.3 |
| Casein free | 3/94 | 3.2 |
| Soy free | 1/94 | 1.1 |
| **Medications Used in Pregnancy** |  |  |
| Zofran | 4/94 | 4.3 |
| Levothyroxine | 4/94 | 4.3 |
| SSRI/SNRI | 3/94 | 3.2 |
| Topiramate | 1/94 | 1.1 |
| Ambien | 1/94 | 1.1 |
| Bupropion | 1/94 | 1.1 |
| Gabapentin | 1/94 | 1.1 |
| Azithromycin | 1/94 | 1.1 |

### Neurobehavioral Profiles

PCA revealed distinct patterns in the distribution of neurobehavioral features across groups. The first two principal components accounted for 40.04% and 13.20% of the total variance, respectively. In the PCA plot, the SKS group showed minimal overlap with healthy controls, with the 95% confidence ellipse for SKS subjects primarily occupying the lower left quadrant of the plot (Fig 1A).

The neurobehavioral profiles of the SKS, PTEN-ASD, and Macrocephaly ASD groups reveal distinct patterns of impairment across multiple domains (Fig 1B). The PTEN-ASD group generally showed moderate difficulties, with elevated scores across motor, social, and executive function measures, though not to the most extreme levels. The Macrocephaly ASD group, while still demonstrating clinically meaningful elevations, generally showed milder impairments across most domains, with the exception of some DCDQ motor scores, social skills as measured by SRS-2, and executive function measures which showed the highest elevations in this group, relative to the other two. The SKS group demonstrated significant impairments across most domains when compared to healthy controls (Table S4). In comparison to PTEN-ASD, there were greater deficits in adaptive and motor functioning (Table S5). Multivariate analysis of variance (MANOVA) revealed significant differences across the three groups in both adaptive functioning and motor abilities, as well as in sensory processing (Table S6). These analyses suggest that while there are some differences within the ASD groups, the SKS group exhibits a particularly broad range of neurobehavioral impairments.

### Correlation of Neurobehavioral Traits Across Protein Domains

Domain-specific analysis of neurobehavioral phenotypes revealed significant differences across MTOR and PTEN protein domains. Kruskal-Wallis tests identified 17 measures showing significant differences between domain groups (p < 0.05), with motor coordination and social behavior showing the strongest effects (Fig 2 and Table S3). The DCDQ Fine Motor measure demonstrated the most significant domain-specific differences (χ² = 20.46, p < 0.001), followed by DCDQ Total Score (χ² = 12.48, p < 0.01) (Fig 2 and Table S3). In both measures, MTOR FAT domain variants were associated with greater impairment compared to PI3K domain variants, though pairwise comparisons within MTOR domains did not reach statistical significance (Tables S3 and S4). In contrast, PTEN domain comparisons showed significant differences across multiple measures, particularly in social responsiveness and executive function domains, with larger sample sizes enabling more robust statistical comparisons (n = 21-42) (Table S4). The phosphatase domain group showed a notably higher representation of individuals with ASD compared to the C2 domain group across all measures examined (Fig 2).

### Correlation of Neurobehavioral Traits with Variant Pathogenicity Scores

Correlation analysis revealed distinct patterns of relationships between missense variant pathogenicity metrics and clinical measures across cohorts (Fig 3). Stability of these correlations was assessed using bootstrap analysis, with “stable” correlations being those categorized as either “Moderately Stable” or “Very Stable”. In the SKS, strong and moderately stable positive correlations were observed, based on bootstrap analysis, between REVEL scores and SSP Auditory Filtering (r=0.77, p < 0.05, uncorrected) and between AlphaMissense scores and SSP Visual/Auditory Sensitivity (r=0.74, p<0.05, uncorrected). A strong and very stable negative correlation was observed between REVEL scores and DCDQ Control During Movement (r=−0.80, p < 0.05, uncorrected). The PTEN cohort showed a very significant stable positive correlation between CADD scores and SSP low energy (r=0.72, p <0.05, uncorrected) and a moderately stable negative correlation with SRS-2 Total T-score (r=−0.64, p<0.05, uncorrected). For the SFARI Genes in the PI3K-AKT-MTOR Cluster cohort, analyses did not reveal any significant and stable correlations based on bootstrap analysis. In the Combined OGID Cohorts analysis (includes SKS, PTEN, and SFARI PI3K cohorts), strong and very stable correlations emerged, based on bootstrap analysis, between CADD scores and multiple behavioral measures. The strongest of these included positive correlations with SSP Low Energy and SSP Total Score and a negative correlation with SRS-2 Total T-Score (all p < 0.05, uncorrected). The strength and direction of correlations varied considerably between cohorts, but the most pronounced and consistently stable relationships, as determined through bootstrap analysis, were observed in the Combined cohorts.

### Diagnostic Classification Using Clinical Decision Trees

We used recursive partitioning to predict membership in diagnostic groups, which produced a stable tree with a relative error of 0.34 and a cross-validated relative error of 0.67 (SD=0.06) (Figure 4A). As the cross-validated relative error is significantly below 1, we have evidence that this decision tree performs above chance in determining group membership. The variables chosen by this algorithm (DCDQ Total Score, SRS-2 Total T-Score, neonatal teeth, VABS-2 Daily Living Skills) jointly explain an estimated 33% of group membership, and specifically of the 18 participants in the MTOR group, 11 are identified correctly by this decision tree, compared to <1 by random guessing. Eight participants are misclassified as MTOR. To determine how much the medical variables contributed to this model, we replicated the analysis with a reduced set of potential predictors, including only behavioral variables. This set of predictors also produced a stable tree with a relative error of 0.51 and a cross-validated relative error of 0.74 (SD=0.06) (Figure 4B). As the cross-validated relative error is significantly below 1, we have evidence that this decision tree performs above chance in determining group membership. The variables chosen by this algorithm (DCDQ Total Score, SRS-2 Total T-Score, DCDQ Fine Motor, ABC Inappropriate Speech, BVMI Standard Score, and ABC Lethargy) jointly explain an estimated 26% of group membership, and specifically, of the 18 participants in the MTOR group, 11 are identified correctly by this decision tree, compared to <1 by random guessing. Only 4 participants are erroneously misclassified as MTOR. The decision tree only performs slightly better by including the medical variables, but the decision tree based on the behavioral data alone performed better at identifying MTOR individuals specifically.

**Figure 4.**
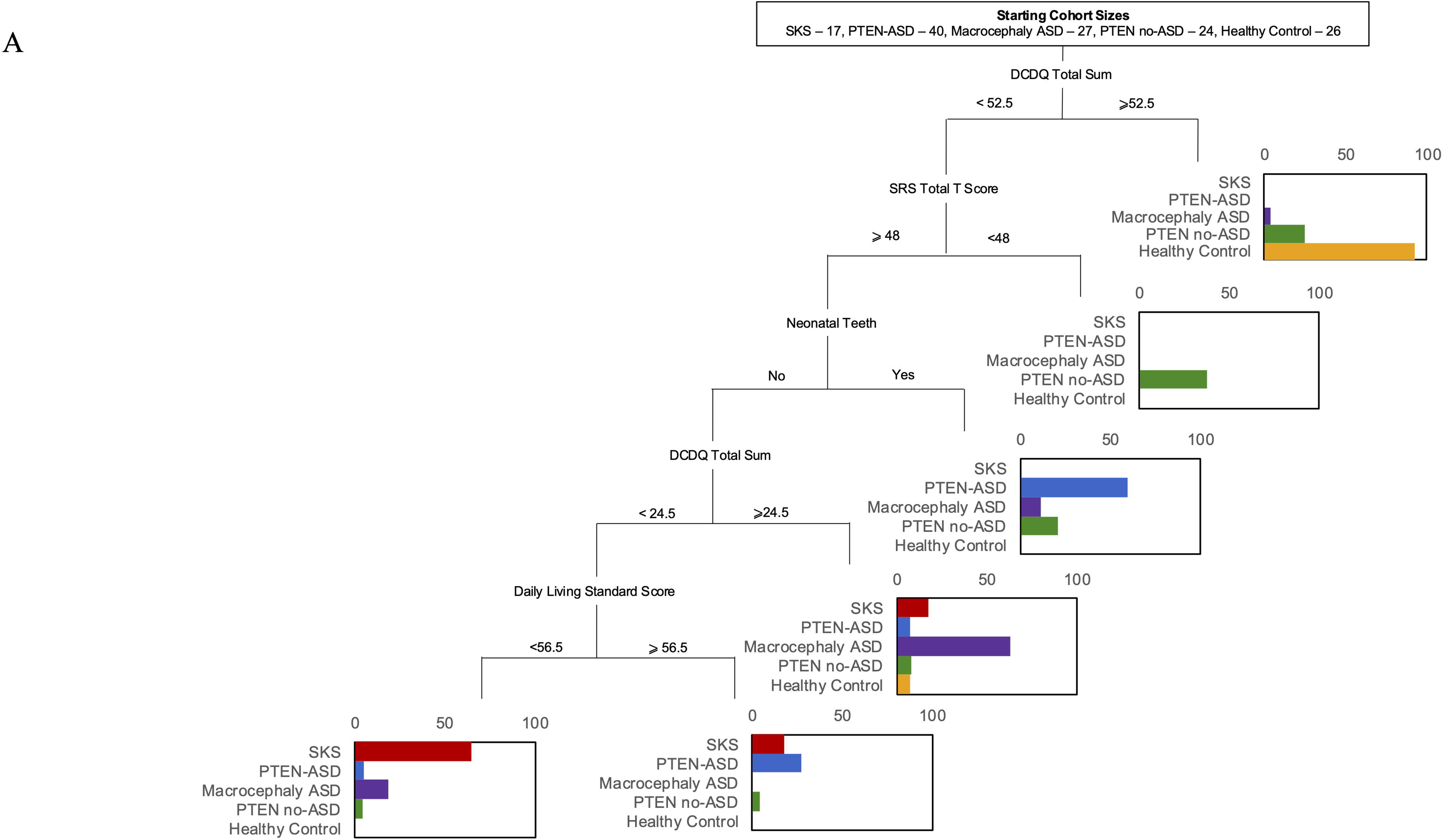
Diagnostic Classification of OGIDs Using Clinical Decision Trees. (A) Decision tree classification model incorporating both behavioral assessments and medical features to differentiate between diagnostic groups (SKS, PTEN-ASD, Macrocephaly ASD, PTEN no-ASD, and Healthy Control). At each branch point, subjects meeting the criterion proceed to the right branch. Bar plots at terminal nodes show the distribution of diagnostic groups, with colors indicating: SKS (red), PTEN-ASD (blue), Macrocephaly ASD (purple), PTEN no-ASD (green), and Healthy Controls (orange). Starting cohort sizes are shown at the top. (B) Decision tree model using behavioral features alone. Both models were developed using recursive partitioning with 10-fold cross-validation to assess generalizability. Abbreviations: DCDQ, Developmental Coordination Disorder Questionnaire; SRS, Social Responsiveness Scale; BVMI, Beery-Buktenica Visual-Motor Integration; ABC, Aberrant Behavior Checklist.

## DISCUSSION

### Novel Phenotypic Findings

We identified novel phenotypic features in SKS, including immune dysregulation and chronic constipation. Immune system abnormalities, though not previously emphasized, align with recent reports indicating potential dysregulation of immune signaling in mTORopathies.^22^ Similarly, constipation has emerged as a significant comorbidity in SKS, which may relate to autonomic dysfunction, gastrointestinal dysmotility or poor diet due to sensory abnormalities. In PHTS, we report a previously undescribed feature present in 44.7% of patients (Table 3)—neonatal teeth— suggesting a broad impact of PTEN mutations on ectodermal derivatives and early developmental processes. Neonatal teeth have been described in other mendelian disorders, including one OGID, Sotos Syndrome. ^23^ In fact, neonatal teeth are one of the features that most strongly distinguishes PHTS (with and without ASD) from SKS in our diagnostic classification (Figure 4A), suggesting it may be an informative clinical marker of PHTS specifically for patients presenting with an OGID phenotype.

### Neurobehavioral Distinctions Between SKS and PHTS

Given that virtually all patients with SKS have ASD and/or ID, we directly compared their neurobehavioral profile to PTEN-ASD and macrocephaly ASD cohorts, using a previously described healthy control cohort^8^ for creation of z-scores. Our findings confirm that individuals with SKS exhibit substantial impairments across neurobehavioral domains compared to individuals without NDDs (Fig 1B). However, impairment is especially pronounced in motor and adaptive functioning, even in comparison to those from the PTEN-ASD and macrocephaly ASD cohorts. Adaptive functioning of SKS individuals, as measured by the VABS-II, was significantly lower than that of PTEN-ASD individuals, particularly in daily living and motor domain scores (Fig 1B; Table S5). All aspects of motor control, as measured on the DCDQ, were also significantly more impaired in SKS individuals than PTEN-ASD individuals. Executive functioning (BRIEF) and emotional and behavioral regulation (CBCL) were similarly preserved compared to PTEN-ASD individuals. There were mixed findings across sensory subdomains, with taste/smell and movement sensitivity significantly less impaired in SKS compared to PTEN-ASD, while all other domains were equally or more impaired. The neurobehavioral profile of SKS may suggest widespread disruption of motor and adaptive functioning circuits, particularly affecting cerebello-cortical pathways, basal ganglia circuits, and frontoparietal networks involved in sensorimotor integration and daily living skills^24^. While executive function networks appear similarly affected as in PTEN-ASD^8^, the distinctive pattern of motor and adaptive impairments in SKS points to a unique impact on neural circuits governing physical coordination and routine behavior execution, potentially reflecting differences in the developmental timing or molecular pathways affected by *MTOR* versus *PTEN* mutations.

### Neurobehavioral Phenotypes Across Protein Domains

Comparative analysis of PTEN domains (Phosphatase vs C2) revealed significant differences across multiple neurobehavioral measures, with the strongest effects observed in social responsiveness (SRS-2; p<0.01) and organizational skills (BRIEF-P; p<0.01), followed by moderate differences in executive function, working memory, and behavioral regulation (p<0.05) (Fig 2; Table S8). In contrast, PTEN domain differences did not significantly impact motor skills, coordination, or adaptive daily living measures. This is consistent with the observation that the majority of individuals with variants in the C2 domain are part of the PTEN-ASD group, while those with variants in the phosphatase domain have on average a milder neurobehavioral phenotype. This is also consistent with previous findings from deep-mutational-scanning of PTEN that shows clinical missense variants cluster most heavily in the phosphatase domain and are depleted in the C2 domain, but with an overall enrichment of gnomAD (non-pathological) variants in the C2 domain.^25^ This suggests that the C2 domain main be more tolerant of minor disruptions such as synonymous or missense variants, but that when more destructive variants are present, such as loss-of-function variants, the neurobehavioral phenotype can be profound, especially for autism behaviors and executive functioning. Notably, no significant differences were observed between MTOR domains (FAT vs PI3K) across any neuropsychological measures (Table S8), though interpretation is limited by smaller sample sizes in the SKS cohort.

### Genotype-Phenotype Correlations and Predictive Challenges

Missense pathogenicity scores including AlphaMissense, CADD, RAS, and REVEL did not consistently correlate with neurobehavioral outcomes in SKS and PHTS cohorts (Table 4). This may have been due to limited power from small sample size, so we combined these cohorts and saw a slightly enhanced stability in correlation measures, as demonstrated via bootstrapping (Fig 3). Overall, CADD scores had the strongest and most stable correlations with neurobehavioral measures across OGIDs, especially sensory measures as measured on the SSP. Auditory filtering was significantly positively correlated with both higher REVEL and CADD pathogenicity scores for SKS, while low energy measures were most strongly correlated to pathogenicity in the PTEN and combined cohorts. Interestingly, sensory profiles derived from the SSP in patients with ASD (not OGIDs specifically) were shown to correlate with CADD scores in clusters of neurodevelopmental genes.^26^ Sensory phenotype subgroups of patients with ASD and/or ID and a known underlying genetic variant (e.g. *ADNP*, *CHD8*, *SCN2A*) have also been identified.^27^ This all suggests that sensory profiles may offer a unique window into underlying genetic pathogenicity, but much larger studies are needed to confirm this.

**Table 4.** Correlation of Neurobehavioral Measures with Variant Pathogenicity Scores Across OGIDs.

|  | **SKS** |  |  |  | **PTEN** |  |  |  | **SFARI PI3K-MTOR-AKT** |  |  | **Combined OGIDs** |  |  |  |
| --- | --- | --- | --- | --- | --- | --- | --- | --- | --- | --- | --- | --- | --- | --- | --- |
|  | AlphaMissense | CADD | RAS | REVEL | AlphaMissense | CADD | RAS | REVEL | AlphaMissense | CADD | REVEL | AlphaMissense | CADD | RAS | REVEL |
| **BRIEF Emot** | -0.12 | -0.29 | -0.13 | -0.37 | 0.27 | -0.3 | -0.05 | 0.06 | - | - | - | 0.06 | -0.31 | -0.09 | -0.32 |
| **BRIEF GEC** | -0.36 | -0.1 | -0.31 | -0.25 | 0.65* | -0.42 | 0.08 | 0.44 | - | - | - | 0.19 | -0.28 | -0.17 | -0.13 |
| **BRIEF Inhib** | -0.53 | -0.26 | -0.52 | -0.51 | 0.48 | -0.41 | 0.01 | 0.21 | - | - | - | -0.04 | -0.33 | -0.36 | -0.32 |
| **BRIEF Shift** | -0.43 | -0.16 | -0.37 | -0.32 | 0.53 | -0.26 | -0.2 | 0.39 | - | - | - | 0.03 | -0.23 | -0.3 | -0.32 |
| **BRIEF WM** | -0.19 | 0.11 | -0.17 | -0.01 | 0.55 | -0.52 | 0.17 | 0.5 | - | - | - | 0.27 | -0.34 | -0.02 | -0.13 |
| **CBCL Ext** | -0.13 | -0.14 | -0.03 | 0.03 | -0.1 | -0.25 | 0 | 0.19 | -0.31 | 0.13 | -0.21 | -0.28* | 0.08 | -0.02 | 0 |
| **CBCL Int** | 0.07 | 0.11 | 0.07 | 0.17 | 0.18 | -0.35 | -0.37 | 0.42 | -0.21 | -0.1 | -0.28 | -0.1 | -0.11 | 0 | 0.04 |
| **CBCL Tot** | -0.19 | -0.1 | -0.1 | -0.06 | 0.19 | -0.2 | -0.34 | 0.38 | -0.26 | 0.02 | -0.26 | -0.21 | -0.02 | -0.14 | 0 |
| **DCDQ Ctrl** | -0.16 | -0.36 | -0.03 | -0.80** | -0.57 | 0.36 | 0.3 | 0.24 | - | - | - | -0.37 | 0.19 | 0.05 | 0.21 |
| **DCDQ Fine** | 0.18 | -0.22 | 0.17 | -0.09 | -0.82** | 0.28 | 0.01 | -0.34 | - | - | - | -0.50* | 0.27 | -0.01 | 0.36 |
| **DCDQ Coord** | -0.29 | -0.22 | -0.29 | -0.47 | -0.52 | 0.29 | 0.23 | -0.23 | - | - | - | -0.34 | 0.22 | -0.07 | 0.33 |
| **DCDQ Tot** | -0.23 | -0.38 | -0.14 | -0.71* | -0.70* | 0.35 | 0.21 | -0.08 | - | - | - | -0.44* | 0.25 | -0.01 | 0.32 |
| **SRS-2 Tot** | -0.3 | -0.41 | -0.18 | -0.3 | 0.52 | -0.64* | 0.08 | 0.52 | - | - | - | 0.2 | -0.58* | -0.04 | -0.33 |
| **SSP Aud** | 0.55* | 0.61* | 0.45 | 0.78** | -0.53 | 0.32 | 0.06 | -0.47 | - | - | - | -0.11 | 0.43* | 0.22 | 0.38 |
| **SSP Energy** | -0.05 | 0.11 | -0.04 | -0.07 | -0.13 | 0.72* | 0.23 | -0.06 | - | - | - | -0.09 | 0.53* | 0.03 | 0.14 |
| **SSP Mvmt** | 0.35 | 0.44 | 0.33 | 0.59* | -0.32 | 0.3 | 0.18 | -0.22 | - | - | - | 0.02 | 0.34 | 0.27 | 0.24 |
| **SSP Tact** | 0.32 | 0.06 | 0.2 | 0.06 | -0.33 | 0.38 | 0.13 | -0.29 | - | - | - | -0.02 | 0.27 | 0.13 | 0.23 |
| **SSP Taste** | 0.25 | 0.33 | 0.14 | 0.34 | -0.27 | 0.27 | 0.22 | -0.28 | - | - | - | -0.01 | 0.3 | 0.14 | 0.12 |
| **SSP Tot** | 0.49 | 0.43 | 0.33 | 0.49 | -0.49 | 0.56* | 0.23 | -0.4 | - | - | - | -0.12 | 0.54* | 0.2 | 0.31 |
| **SSP Under** | 0.02 | 0.28 | -0.14 | 0.08 | -0.4 | 0.3 | 0.26 | -0.33 | - | - | - | -0.25 | 0.31 | 0.01 | 0.11 |
| **SSP Vis** | 0.51 | 0.09 | 0.36 | 0.39 | -0.49 | 0.37 | -0.1 | -0.28 | - | - | - | 0.1 | 0.22 | 0.25 | 0.21 |
| **VABS ABC** | 0.02 | -0.45 | 0.14 | -0.23 | -0.87** | 0.45 | -0.14 | -0.64* | -0.24 | 0.14 | -0.65 | -0.29 | 0.01 | 0.06 | 0.09 |
| **VABS Comm** | 0.02 | -0.51 | 0.2 | -0.28 | -0.75* | 0.43 | -0.11 | -0.55 | -0.21 | 0.25 | -0.46 | -0.28 | 0.06 | 0.07 | 0.21 |
| **VABS DLS** | 0.01 | -0.45 | 0.13 | -0.29 | -0.89** | 0.44 | -0.11 | -0.63* | -0.18 | 0.14 | -0.52 | -0.28 | 0.03 | 0.06 | 0.08 |
| **VABS Motor** | 0.2 | -0.46 | 0.48 | -0.44 | -0.4 | -0.41 | -0.33 | -0.2 | 0.11 | 0.15 | - | 0.02 | -0.19 | 0.18 | 0.11 |
| **VABS Soc** | 0.16 | -0.34 | 0.22 | -0.11 | -0.77* | 0.52 | -0.19 | -0.64* | -0.35 | 0.01 | -0.38 | -0.29 | -0.01 | 0.07 | 0.21 |

SKS = Smith-Kingsmore Syndrome; PTEN = Phosphatase and Tensin Homolog gene; SFARI PI3K-MTOR-AKT = Simons Foundation Autism Research Initiative PI3K-MTOR-AKT pathway genes; OGIDs = Overgrowth Intellectual Disability Syndromes. Variant impact predictions were calculated using AlphaMissense, CADD, RAS, and REVEL. Behavioral measures include BRIEF (Behavior Rating Inventory of Executive Function) subscales: Emotional Control (Emot), Global Executive Composite (GEC), Inhibition (Inhib), Cognitive Flexibility (Shift), and Working Memory (WM); CBCL (Child Behavior Checklist) scores: Externalizing (Ext), Internalizing (Int), and Total (Tot); DCDQ (Developmental Coordination Disorder Questionnaire) domains: Control During Movement (Ctrl), Fine Motor Control (Fine), General Coordination (Coord), and Total Score (Tot); SRS-2 (Social Responsiveness Scale-2) Total Score; SSP (Short Sensory Profile) subscales: Auditory Processing (Aud), Low Energy/Weak (Energy), Movement Sensitivity (Mvmt), Tactile Sensitivity (Tact), Taste/Smell Sensitivity (Taste), Total Score (Tot), Underresponsive/Seeks Sensation (Under), and Visual/Auditory Sensitivity (Vis); and VABS (Vineland Adaptive Behavior Scales) domains: Adaptive Behavior Composite (ABC), Communication (Comm), Daily Living Skills (DLS), Motor Skills (Motor), and Socialization (Soc). \* $p < 0.05$ , \*\* $p\text{FDR} < 0.05$ , - = missing data or not applicable.

### Diagnostic Classification of OGIDs Using Clinical Decision Trees

We explored the potential for clinical decision trees to aid in diagnostic classification of OGIDs. The resulting decision trees, generated using recursive partitioning, highlight the relative contributions of specific behavioral and medical features in distinguishing between diagnostic groups. The observation that a model incorporating both behavioral assessments and the presence of neonatal teeth performed above chance suggests that a combined approach leveraging both clinical and developmental characteristics may enhance diagnostic accuracy. The inclusion of neonatal teeth, a feature particularly prominent in the PHTS cohort, as a distinguishing feature in the diagnostic classification emphasizes the significance of even seemingly minor physical findings in differentiating OGID subtypes. This underscores the need for careful consideration of seemingly subtle clinical features alongside detailed neurodevelopmental assessments.

However, the finding that a decision tree based solely on neurobehavioral assessments demonstrated a similar, albeit slightly reduced, capacity to predict diagnostic group membership carries significant clinical implications. This suggests that a comprehensive neurobehavioral evaluation alone may offer substantial information in guiding initial diagnostic considerations, particularly when genetic testing results or a complete medical history are not immediately available. The identified behavioral features, such as measures of motor coordination, social communication, and executive function, therefore act as key variables in driving diagnostic classification for clinicians.

The limited variance explained by both decision tree models further underscores the multifactorial nature of these disorders and implies that prediction of clinical outcomes must account for a more complex network of genetic, environmental, and developmental factors. The fact that these models still perform above change however lends promise to using these tools as a starting point, and underscores the value of larger more complex and potentially personalized diagnostic and interventional strategies. Future work integrating longitudinal neurobehavioral data, genetic variants, and potentially neuroimaging findings may allow for the creation of more robust and accurate predictive models, further guiding diagnostic evaluations and the development of targeted intervention strategies for individuals with OGIDs.

### Limitations

Our study is limited by its cross-sectional design and relatively small sample size, particularly for SKS, which restricts the generalizability of our findings. Additionally, while we utilized standardized neurobehavioral assessments, potential biases introduced by parent-report measures should be considered. Future studies incorporating objective neurophysiological and neuroimaging data could provide more mechanistic insights into the observed neurobehavioral patterns.

## Conclusion

This study enhances our understanding of SKS by providing a detailed neurobehavioral profile and establishing key distinctions from PHTS. While both conditions share core impairments in adaptive functioning and executive control, SKS is marked by particularly severe motor deficits and adaptive functioning challenges. The significantly lower VABS-II daily living and motor domain scores in SKS highlight broader impairments in functional independence that necessitate tailored intervention approaches. The identification of immune dysfunction and gastrointestinal involvement in SKS, along with neonatal teeth in PTEN-related conditions, expands the clinical spectrum of these OGIDs and warrants further investigation. The lack of robust genotype-phenotype correlations underscores the need for individualized clinical assessments rather than reliance on variant pathogenicity scores for prognostication, although further investigation into the link between sensory phenotypes and genetic pathogenicity subtypes is warranted. Moving forward, longitudinal studies following the developmental trajectory of neurobehavioral features will be critical, as will expanding comparative analyses to include additional OGIDs. This will require coordinated efforts across patient advocacy groups and research centers to gather sufficient data on these rare conditions. Future research should also incorporate neuroimaging and neurophysiological measures to better understand the mechanisms underlying the distinct patterns of impairment observed in different PI3K-AKT-MTOR pathway disorders.

## DATA AVAILABILITY

Anonymized data will be shared by request from a qualified academic investigator.

## ACKNOWLEDGEMENTS

We are also sincerely indebted to the generosity of the Smith-Kingsmore Syndrome families and patients and those in PTEN clinics across the United States who contributed their time and effort to this study. We would also like to thank the PTEN Hamartoma Syndrome Foundation and the PTEN Research for their continued support in PTEN research and the Smith-Kingsmore Syndrome Foundation.

## The Developmental Synaptopathies Consortium (DSC) – PTEN Hamartoma

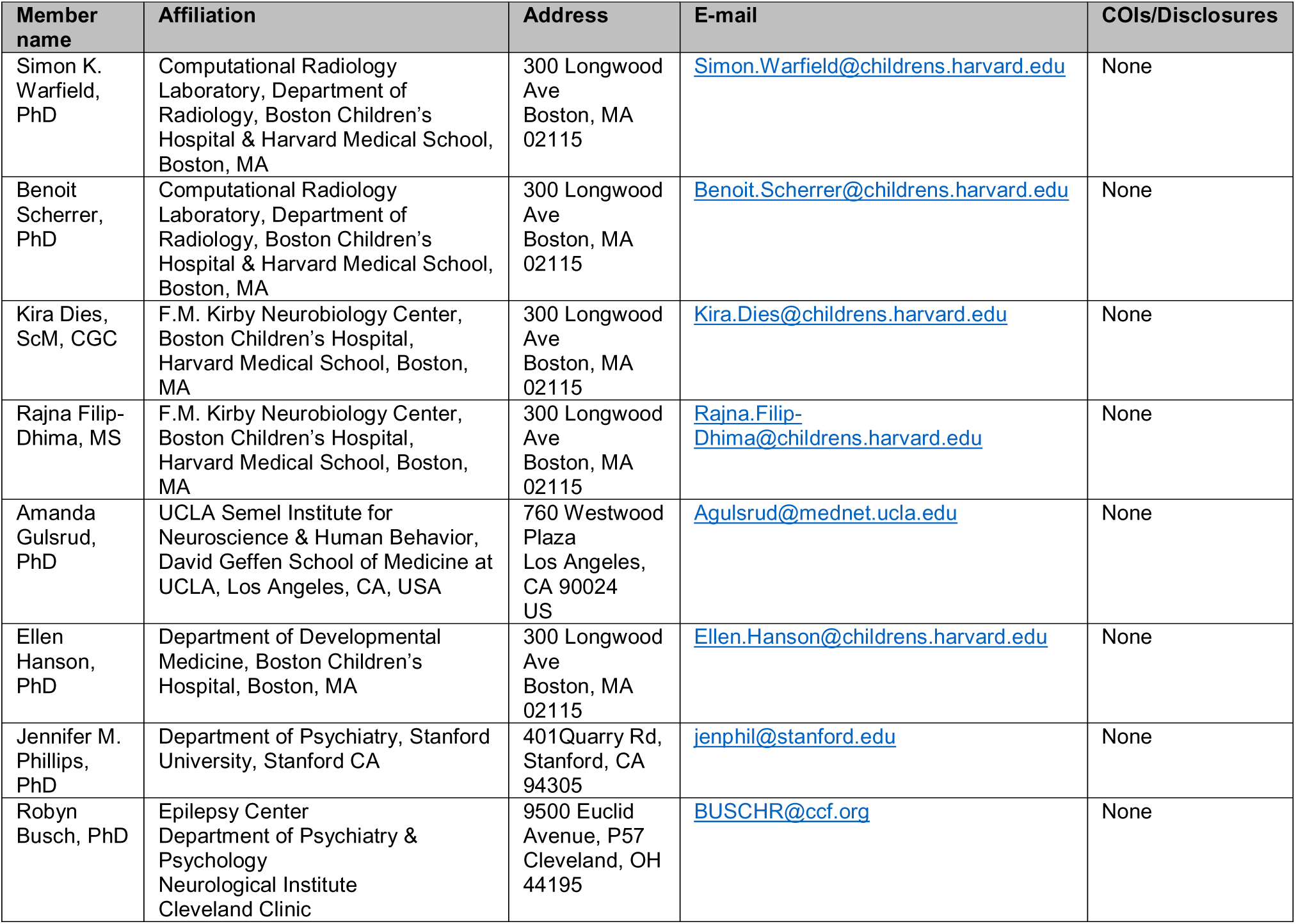

## Syndrome Group

## SUPPLEMENTARY MATERIAL

Supplementary material is available at Brain online

