## Supplement for "Neurobehavioral Signatures in Overgrowth Intellectual Disability Syndromes: Dissecting Genotype-Phenotype Relationships in the PI3K-AKT-MTOR Pathway"

**Table S1.** Neurobehavioral Battery Adapted from the PHTS Developmental Synaptopathies Consortium

| Measure | Domain |
|---|---|
| Social Responsiveness Scale 2 (SRS-2) | ASD-like Behaviors |
| Repetitive Behavioral Scale Revised (RBS-R) | Repetitive Behaviors |
| Child Behavior Checklist Adult/Observer Report Form (CBCL/ARF) | Behavioral Functioning |
| Aberrant Behavior Checklist – Community (ABC-C)[a] | Aberrant Behaviors |
| Vineland Adaptive Behavior Scale (VABS-II) | Adaptive Functioning |
| Developmental Coordination Disorder Questionnaire (DCDQ) | Motor Functioning |
| Behavior Rating Inventory of Executive Function (BRIEF) | Executive Functioning |
| Short Sensory Profile (SSP) | Sensory Functioning |

[a] Not included in original DSC battery

**Table S2.** Descriptive Characteristics of Study Cohorts

| Cohort | N | Median Age (Months) | Age IQR | Female Proportion |
|---|---|---|---|---|
| Healthy Control | 30 | 156 | 105-207.8 | 0.53 |
| SKS | 17 | 57 | 45-97 | 0.24 |
| Macrocephaly ASD | 32 | 126 | 89.3-204.8 | 0.16 |
| PTEN no-ASD | 28 | 104 | 67.75-132 | 0.36 |
| PTEN-ASD | 43 | 107 | 66-140.5 | 0.21 |

IQR = Interquartile range; N = number of participants

**Table S3.** Neurobehavioral Profile of SKS and PTEN Patients

| Measure | SKS median | SKS IQR | PTEN ASD median | PTEN ASD IQR | Macrocephaly ASD median | Macrocephaly ASD IQR | PTEN No-ASD Median | PTEN no-ASD IQR | Healthy Control median | Healthy Control IQR |
|---|---|---|---|---|---|---|---|---|---|---|
| ABC Hyperactivity | 12 | 8.8-15.2 | 9 | 6.0-16.8 | 13 | 8.5-19.5 | 4 | 1.0-9.2 | NA | NA |
| ABC Irritability | 11 | 7.0-15.2 | 5 | 1.2-11.0 | 10 | 4.0-13.0 | 1 | 0.0-5.2 | NA | NA |
| ABC Lethargy | 9.5 | 4.0-12.0 | 7.5 | 2.0-18.0 | 10 | 7.5-14.5 | 1 | 0.0-5.5 | NA | NA |
| ABC Speech | 1 | 0.0-3.2 | 3 | 2.0-5.0 | 3 | 2.0-4.0 | 1 | 0.0-2.2 | NA | NA |
| ABC Stereotypies | 7 | 3.0-9.5 | 4.5 | 1.0-8.0 | 5 | 2.0-8.5 | 0 | 0.0-1.2 | NA | NA |
| ABC Total Score | 42 | 31.8-53.8 | 32 | 19.2-57.2 | 39 | 33.5-50.5 | 13 | 5.0-22.8 | NA | NA |
| BRIEF Emotional Control | 56 | 55.0-64.0 | 57 | 50.5-64.2 | 64 | 55.5-73.2 | 50.5 | 39.5-57.2 | 44.5 | 37.0-50.2 |
| BRIEF Inhibit | 58 | 52.0-63.0 | 60 | 54.5-69.2 | 69 | 62.2-72.8 | 46.5 | 44.8-57.8 | 45 | 41.0-49.0 |
| BRIEF Plan/Organize | 57 | 46.0-68.0 | 65.5 | 51.8-76.0 | 71.5 | 61.8-75.0 | 51 | 42.5-70.0 | 42 | 38.8-58.8 |
| BRIEF Shift | 60 | 45.0-69.0 | 62.5 | 49.8-70.2 | 68 | 59.8-74.0 | 53 | 45.2-56.0 | 40.5 | 38.8-49.2 |
| BRIEF Total | 63 | 56.0-73.0 | 66 | 56.5-76.0 | 73 | 65.2-78.0 | 53 | 42.0-73.5 | 42.5 | 37.8-53.2 |
| BRIEF Working Memory | 68 | 66.0-74.0 | 73 | 59.5-78.5 | 71.5 | 62.2-79.8 | 56.5 | 47.2-75.2 | 45 | 40.0-49.2 |
| CBCL Externalizing | 57 | 47.5-64.0 | 53.5 | 43.0-60.0 | 58 | 48.0-63.8 | 41 | 36.5-43.5 | 48.5 | 34.0-53.2 |
| CBCL Internalizing | 59 | 57.2-64.2 | 63 | 55.0-67.2 | 61.5 | 52.0-67.5 | 48 | 37.5-63.5 | 48 | 34.0-55.2 |
| CBCL Total | 60.5 | 58.0-67.2 | 63.5 | 52.5-71.0 | 66.5 | 59.2-70.0 | 46 | 44.5-60.5 | 45.5 | 34.0-53.2 |
| DCDQ Control | 6 | 6.0-9.0 | 9 | 8.0-13.0 | 14.5 | 10.2-15.8 | 14 | 10.0-23.0 | 30 | 25.5-30.0 |
| DCDQ Coordination | 7 | 5.0-8.0 | 8 | 6.0-11.0 | 10 | 7.2-12.0 | 14 | 11.0-18.5 | 22 | 18.5-25.0 |
| DCDQ Fine Motor | 4 | 4.0-4.0 | 5 | 4.0-8.0 | 7.5 | 4.2-12.8 | 10 | 7.0-14.0 | 20 | 18.5-20.0 |
| DCDQ Total Score | 17.5 | 15.0-20.8 | 23 | 21.0-34.0 | 33.5 | 25.5-41.5 | 38 | 27.5-54.0 | 70 | 64.0-75.0 |
| RBS-R Total Score | 19 | 14.5-38.0 | 20.5 | 11.0-46.5 | 26.5 | 15.5-33.2 | 3.5 | 1.0-15.2 | NA | NA |
| SRS-2 Total Score | 75 | 70.0-85.0 | 75.5 | 68.5-83.0 | 77 | 71.0-81.8 | 47 | 44.0-65.0 | 43 | 40.5-47.0 |
| SSP Auditory Filtering | 18 | 15.8-20.0 | 18 | 14.2-21.5 | 18 | 16.0-19.8 | 23 | 20.0-27.0 | 28 | 23.8-30.0 |
| SSP Low Energy | 15.5 | 14.0-18.2 | 15 | 8.0-21.8 | 23 | 16.5-28.0 | 20 | 14.8-27.5 | 30 | 29.0-30.0 |
| SSP Movement | 14.5 | 11.0-15.0 | 11.5 | 9.0-15.0 | 14 | 12.0-15.0 | 13 | 10.0-15.0 | 15 | 14.0-15.0 |

| | | | | | | | | | | |
|---|---|---|---|---|---|---|---|---|---|---|
| SSP Tactile | 27.5 | 23.0-29.3 | 28 | 24.0-31.8 | 28.5 | 27.0-32.0 | 32 | 30.8-35.0 | 35 | 33.0-35.0 |
| SSP Taste/Smell | 18.5 | 11.8-20.0 | 16 | 7.2-20.0 | 17.5 | 12.0-20.0 | 19 | 15.8-20.0 | 20 | 16.8-20.0 |
| SSP Total | 124 | 111.4-136.0 | 120 | 102.8-140.8 | 140 | 126.0-149.0 | 154.5 | 134.8-172.8 | 185 | 163.0-188.5 |
| SSP Underresponsive/Seeks | 20 | 16.0-22.0 | 20.5 | 15.5-27.0 | 22.5 | 19.2-27.5 | 31.5 | 22.8-34.2 | 35 | 29.8-35.0 |
| SSP Visual/Auditory | 16 | 13.0-20.2 | 16 | 13.0-19.0 | 18 | 14.2-20.8 | 21 | 17.8-24.0 | 25 | 18.0-25.0 |
| VABS-II Adaptive Behavior | 53.5 | 46.2-59.8 | 62 | 55.0-75.2 | 61 | 55.0-70.0 | 95.5 | 80.2-111.8 | 94 | 88.5-109.8 |
| VABS-II Communication | 59 | 42.0-65.0 | 64 | 56.0-78.0 | 66 | 57.5-74.2 | 92.5 | 83.5-113.5 | 100 | 91.2-111.0 |
| VABS-II Daily Living | 52 | 46.0-56.0 | 66 | 58.0-77.0 | 66 | 56.5-73.0 | 101 | 81.5-109.8 | 96 | 87.5-106.5 |
| VABS-II Motor | 51 | 37.0-61.0 | 68.5 | 61.0-75.8 | 67 | 61.0-91.0 | 78 | 70.0-91.0 | 111 | 102.0-115.5 |
| VABS-II Socialization | 61 | 57.0-68.0 | 68.5 | 56.5-76.8 | 59 | 48.0-69.0 | 104.5 | 88.0-119.0 | 94.5 | 85.0-103.0 |

Values shown as median (interquartile range). SKS: MTOR-associated Smithsonian-Kingsmore Syndrome; PTEN-ASD: PTEN-associated autism spectrum disorder. Measures: SRS-2 (Social Responsiveness Scale-2; higher scores indicate greater social impairment), RBS-R (Repetitive Behavior Scale-Revised; higher scores indicate more repetitive behaviors), VABS-II (Vineland Adaptive Behavior Scales-II; lower scores indicate greater impairment), CBCL (Child Behavior Checklist; higher scores indicate more behavioral problems), DCDQ (Developmental Coordination Disorder Questionnaire; lower scores indicate greater motor impairment), ABC (Aberrant Behavior Checklist; higher scores indicate more problematic behaviors), BRIEF (Behavior Rating Inventory of Executive Function; higher scores indicate greater executive function difficulties), SSP (Short Sensory Profile; lower scores indicate greater sensory differences).

**Table S4.** Neurobehavioral Feature Comparison Between SKS and Healthy Controls

| Domain | Measure | N SKS | N Healthy Control | W | p-value | Effect Size (r) | p-value adj | Sig | Sig adj |
|---|---|---|---|---|---|---|---|---|---|
| Adaptive Function | VABS-II Adaptive Behavior | 22 | 10 | 198 | 3.70E-04 | 0.63 | 4.60E-04 | *** | *** |
| Adaptive Function | VABS-II Communication | 22 | 13 | 270 | 1.50E-05 | 0.73 | 7.70E-05 | *** | *** |
| Adaptive Function | VABS-II Daily Living | 22 | 12 | 241 | 9.10E-05 | 0.67 | 2.30E-04 | *** | *** |
| Adaptive Function | VABS-II Motor | 15 | 9 | 120 | 0.002 | 0.63 | 0.002 | ** | ** |
| Adaptive Function | VABS-II Socialization | 22 | 11 | 220.5 | 1.50E-04 | 0.66 | 2.60E-04 | *** | *** |
| Autism Symptoms | SRS-2 Total Score | 20 | 16 | 1.5 | 4.80E-07 | 0.84 | 4.80E-07 | *** | *** |
| Behavior | CBCL Externalizing | 24 | 16 | 96 | 0.008 | 0.42 | 0.008 | ** | ** |
| Behavior | CBCL Internalizing | 24 | 16 | 73 | 0.001 | 0.52 | 0.002 | ** | ** |
| Behavior | CBCL Total | 24 | 16 | 48 | 7.30E-05 | 0.63 | 2.20E-04 | *** | *** |
| Executive Function | BRIEF Emotional Control | 24 | 13 | 77.5 | 0.013 | 0.41 | 0.015 | * | * |
| Executive Function | BRIEF Inhibit | 24 | 13 | 59 | 0.002 | 0.51 | 0.003 | ** | ** |
| Executive Function | BRIEF Plan/Organize | 24 | 13 | 99 | 0.072 | 0.3 | 0.072 | | |
| Executive Function | BRIEF Shift | 24 | 13 | 60 | 0.002 | 0.5 | 0.003 | ** | ** |
| Executive Function | BRIEF Total | 24 | 13 | 55 | 0.001 | 0.53 | 0.003 | ** | ** |
| Executive Function | BRIEF Working Memory | 24 | 13 | 27 | 4.10E-05 | 0.67 | 2.50E-04 | *** | *** |
| Motor | DCDQ Control | 23 | 16 | 366.5 | 7.10E-08 | 0.86 | 1.40E-07 | *** | *** |
| Motor | DCDQ Coordination | 23 | 16 | 368 | 1.10E-07 | 0.85 | 1.40E-07 | *** | *** |
| Motor | DCDQ Fine Motor | 23 | 16 | 368 | 3.20E-08 | 0.89 | 1.30E-07 | *** | *** |
| Motor | DCDQ Total Score | 23 | 16 | 368 | 1.40E-07 | 0.84 | 1.40E-07 | *** | *** |
| Sensory | SSP Auditory Filtering | 24 | 16 | 344.5 | 2.20E-05 | 0.67 | 3.50E-05 | *** | *** |
| Sensory | SSP Low Energy | 24 | 16 | 384 | 3.60E-08 | 0.87 | 2.90E-07 | *** | *** |
| Sensory | SSP Movement | 24 | 16 | 241 | 0.123 | 0.24 | 0.123 | | |
| Sensory | SSP Tactile | 24 | 16 | 345.5 | 1.70E-05 | 0.68 | 3.30E-05 | *** | *** |
| Sensory | SSP Taste/Smell | 24 | 16 | 246 | 0.101 | 0.26 | 0.115 | | |
| Sensory | SSP Total | 24 | 15 | 354 | 5.20E-07 | 0.8 | 2.10E-06 | *** | *** |
| Sensory | SSP Underresponsive/Seeks | 24 | 15 | 338 | 3.50E-06 | 0.74 | 9.40E-06 | *** | *** |

| | | | | | | | | | |
|---|---|---|---|---|---|---|---|---|---|
| Sensory | SSP Visual/Auditory | 24 | 16 | 322 | 2.50E-04 | 0.58 | 3.40E-04 | *** | *** |

Mann-Whitney U tests were performed to compare behavioral measures between individuals with MTOR-associated Smithsonian-Kingsmore Syndrome (SKS) and healthy controls, with false discovery rate adjustment (adj) correction applied within each domain. N SKS and N Healthy Controls indicate number of subjects with data available for each measure. W: Mann-Whitney U test statistic; r: effect size. Measures: SRS-2 (Social Responsiveness Scale-2), RBS-R (Repetitive Behavior Scale-Revised), VABS-II (Vineland Adaptive Behavior Scales-II), CBCL (Child Behavior Checklist), DCDQ (Developmental Coordination Disorder Questionnaire), ABC (Aberrant Behavior Checklist), BRIEF (Behavior Rating Inventory of Executive Function), SSP (Short Sensory Profile). *p<0.05, **p<0.01, ***p<0.001.

**Table S5.** Neurobehavioral Feature Comparison Between SKS and PTEN-ASD

| Domain | Measure | N SKS | N PTEN-ASD | W | p-value | Effect size (r) | p-value adj | Sig | Sig adj |
|---|---|---|---|---|---|---|---|---|---|
| Aberrant Behavior | ABC Hyperactivity | 16 | 38 | 356 | 0.328 | 0.13 | 0.418 | | |
| Aberrant Behavior | ABC Irritability | 16 | 38 | 404 | 0.059 | 0.26 | 0.176 | | |
| Aberrant Behavior | ABC Lethargy | 16 | 38 | 299 | 0.932 | 0.01 | 0.932 | | |
| Aberrant Behavior | ABC Speech | 16 | 38 | 199 | 0.044 | 0.27 | 0.176 | * | |
| Aberrant Behavior | ABC Stereotypies | 16 | 38 | 377 | 0.168 | 0.19 | 0.336 | | |
| Aberrant Behavior | ABC Total Score | 16 | 38 | 354 | 0.348 | 0.13 | 0.418 | | |
| Adaptive Function | VABS-II Adaptive Behavior | 10 | 36 | 105 | 0.047 | 0.29 | 0.079 | * | |
| Adaptive Function | VABS-II Communication | 13 | 37 | 165 | 0.097 | 0.23 | 0.121 | | |
| Adaptive Function | VABS-II Daily Living | 12 | 37 | 108 | 0.008 | 0.38 | 0.041 | ** | * |
| Adaptive Function | VABS-II Motor | 9 | 18 | 38.5 | 0.03 | 0.42 | 0.076 | * | |
| Adaptive Function | VABS-II Socialization | 11 | 36 | 172.5 | 0.53 | 0.09 | 0.53 | | |
| Autism Symptoms | RBS-R Total Score | 16 | 38 | 303 | 0.992 | 0 | 0.992 | | |
| Autism Symptoms | SRS-2 Total Score | 16 | 38 | 349 | 0.399 | 0.11 | 0.797 | | |
| Behavior | CBCL Externalizing | 16 | 16 | 155 | 0.317 | 0.18 | 0.952 | | |
| Behavior | CBCL Internalizing | 16 | 16 | 120.5 | 0.792 | 0.05 | 0.97 | | |
| Behavior | CBCL Total | 16 | 16 | 129.5 | 0.97 | 0.01 | 0.97 | | |
| Executive Function | BRIEF Emotional Control | 13 | 32 | 208 | 1 | 0 | 1 | | |
| Executive Function | BRIEF Inhibit | 13 | 32 | 165 | 0.286 | 0.16 | 0.859 | | |
| Executive Function | BRIEF Plan/Organize | 13 | 32 | 153 | 0.172 | 0.2 | 0.859 | | |
| Executive Function | BRIEF Shift | 13 | 32 | 185 | 0.573 | 0.08 | 0.87 | | |
| Executive Function | BRIEF Total | 13 | 31 | 179.5 | 0.58 | 0.08 | 0.87 | | |
| Executive Function | BRIEF Working Memory | 13 | 32 | 194.5 | 0.744 | 0.05 | 0.893 | | |
| Motor | DCDQ Control | 16 | 37 | 160.5 | 0.008 | 0.36 | 0.011 | ** | * |
| Motor | DCDQ Coordination | 16 | 37 | 192 | 0.042 | 0.28 | 0.042 | * | * |
| Motor | DCDQ Fine Motor | 16 | 37 | 106 | 7.70E-05 | 0.54 | 3.10E-04 | *** | *** |
| Motor | DCDQ Total Score | 16 | 37 | 125.5 | 9.50E-04 | 0.45 | 0.002 | *** | ** |

| | | | | | | | |
|---|---|---|---|---|---|---|---|
| Sensory | SSP Auditory Filtering | 16 | 38 | 301.5 | 0.97 | 0.01 | 0.97 |
| Sensory | SSP Low Energy | 16 | 38 | 319 | 0.783 | 0.04 | 0.97 |
| Sensory | SSP Movement | 16 | 38 | 396.5 | 0.074 | 0.24 | 0.592 |
| Sensory | SSP Tactile | 16 | 38 | 280 | 0.656 | 0.06 | 0.97 |
| Sensory | SSP Taste/Smell | 16 | 38 | 374.5 | 0.175 | 0.18 | 0.699 |
| Sensory | SSP Total | 15 | 38 | 291 | 0.913 | 0.01 | 0.97 |
| Sensory | SSP Underresponsive/Seeks | 15 | 38 | 237.5 | 0.353 | 0.13 | 0.941 |
| Sensory | SSP Visual/Auditory | 16 | 38 | 320.5 | 0.761 | 0.04 | 0.97 |

Mann-Whitney U tests were performed to compare behavioral measures between individuals with SKS and PTEN-associated autism spectrum disorder (PTEN-ASD), with FDR correction applied within each domain (adj). N SKS and N PTEN-ASD indicate number of subjects with data available for each measure. r: effect size; W: Mann-Whitney U test statistic;. Domain measures as defined in Table 1. *$p<0.05$, **$p<0.01$, ***$p<0.001$

**Table S6.** MANOVA results comparing SKS, PTEN-ASD, and Macrocephaly ASD groups

| Domain | Total N | Pillai | F | df | p-value | Sig |
|---|---|---|---|---|---|---|
| Adaptive Function | 40 | 0.47 | 2.1 | 10, 68 | 0.036 | * |
| Autism Symptoms | 79 | 0.06 | 1.19 | 4, 152 | 0.317 | |
| Behavior | 50 | 0.14 | 1.13 | 6, 92 | 0.35 | |
| Executive Function | 68 | 0.22 | 1.25 | 12, 122 | 0.257 | |
| Motor | 79 | 0.31 | 3.43 | 8, 148 | 0.001 | ** |
| Sensory | 78 | 0.33 | 1.73 | 16, 138 | 0.047 | * |

Multivariate analysis of variance (MANOVA) was performed to compare behavioral measures across SKS, PTEN-ASD, and macrocephaly-associated ASD groups. N indicates number of subjects with complete data for all measures within each domain. Pillai: Pillai's trace statistic; F: F-statistic; df: degrees of freedom. Domain measures as defined in Table 1. *p<0.05, **p<0.01, ***p<0.001.

**Table S7.** Kruskal-Wallis Test Results for Neuropsychological Measures Across MTOR/PTEN Domains

| Phenotype | Chi-squared | df | p-value | Significance |
|---|---|---|---|---|
| DCDQ Fine Motor | 20.461118 | 3 | 0.000136 | *** |
| DCDQ Total Score | 12.483682 | 3 | 0.005897 | ** |
| SRS-2 Total T-Score | 11.328336 | 3 | 0.010077 | * |
| VABS-2 Daily Living Skills | 11.251485 | 3 | 0.010441 | * |
| ABC Irritability | 9.911848 | 3 | 0.019331 | * |
| RBSR Total Score | 9.671468 | 3 | 0.021575 | * |
| CBCL Externalizing | 9.506216 | 3 | 0.023265 | * |
| ABC Stereotypy | 9.252974 | 3 | 0.02611 | * |
| BRIEF-P Emotional Control | 8.945154 | 3 | 0.030029 | * |
| VABS-2 Communication | 8.881718 | 3 | 0.030906 | * |
| CBCL Total | 8.69116 | 3 | 0.033692 | * |
| VABS-2 Adaptive Behavior Composite | 8.509843 | 3 | 0.03657 | * |
| BRIEF-P Working Memory | 8.444515 | 3 | 0.037665 | * |
| DCDQ Control During Movement | 8.354609 | 3 | 0.039224 | * |
| DCDQ General Coordination | 8.286289 | 3 | 0.040451 | * |
| BRIEF-P Organization of Materials | 7.87418 | 3 | 0.048685 | * |
| ABC Total Score | 7.834389 | 3 | 0.049561 | * |
| CBCL Internalizing | 7.160743 | 3 | 0.066947 | ns |
| BRIEF-P Global Executive Composite | 6.329578 | 3 | 0.096631 | ns |
| BRIEF-P Behavioral Regulation Index | 6.256085 | 3 | 0.099795 | ns |
| VABS-2 Motor Skills | 6.225118 | 3 | 0.101157 | ns |
| BRIEF-P Initiate | 6.223361 | 3 | 0.101235 | ns |
| ABC Lethargy | 6.038411 | 3 | 0.109756 | ns |
| ABC Hyperactivity | 4.713158 | 3 | 0.194047 | ns |
| VABS-2 Socialization | 4.623689 | 3 | 0.201519 | ns |
| BRIEF-P Metacognition Index | 4.546817 | 3 | 0.208152 | ns |
| ABC Inappropriate Speech | 4.482657 | 3 | 0.213842 | ns |
| BRIEF-P Shift | 4.481134 | 3 | 0.213979 | ns |
| BRIEF-P Plan/Organize | 3.618187 | 3 | 0.305754 | ns |
| BRIEF-P Inhibition | 3.221903 | 3 | 0.358661 | ns |
| BVMI Standard Score | 1.028643 | 2 | 0.597906 | ns |
| BRIEF-P Monitor | 1.843478 | 3 | 0.60552 | ns |

The table presents the results of Kruskal-Wallis tests performed to compare different protein domains across various neuropsychological measures. Significance levels are indicated as follows: * $p<0.05$, ** $p<0.01$, *** $p<0.001$, ns not significant. **Abbreviations**: VABS, Vineland Adaptive Behavior Scales; SRS, Social Responsiveness Scale; CBCL, Child Behavior Checklist; BRIEF-P, Behavior Rating Inventory of Executive Function - Preschool Version; ABC, Aberrant Behavior Checklist; DCDQ, Developmental Coordination Disorder Questionnaire; BVMI, Beery-Buktenica Developmental Test; df, degrees of freedom.

**Table S8.** Pairwise Comparison Results for Neuropsychological Measures Across MTOR/PTEN Domains

| Phenotype | PTEN p-value | PTEN N | MTOR p-value | MTOR N | PTEN Sig | MTOR Sig |
|---|---|---|---|---|---|---|
| SRS-2 Total T-Score | 0.007596 | 39 | 0.828139 | 15 | ** | ns |
| BRIEF-P Organization of Materials | 0.009822 | 30 | 1 | 5 | ** | ns |
| RBSR Total Score | 0.01089 | 41 | 0.47009 | 15 | * | ns |
| BRIEF-P Working Memory | 0.017942 | 31 | 1 | 5 | * | ns |
| CBCL Internalizing | 0.020323 | 21 | 1 | 15 | * | ns |
| CBCL Total | 0.022851 | 21 | 0.217806 | 15 | * | ns |
| BRIEF-P Initiate | 0.03487 | 30 | 0.723674 | 5 | * | ns |
| BRIEF-P Global Executive Composite | 0.041655 | 30 | 0.723674 | 5 | * | ns |
| BRIEF-P Emotional Control | 0.044027 | 31 | 0.723674 | 5 | * | ns |
| BRIEF-P Metacognition Index | 0.049685 | 30 | 1 | 5 | * | ns |
| ABC Inappropriate Speech | 0.066253 | 42 | 0.764503 | 15 | ns | ns |
| CBCL Externalizing | 0.081317 | 21 | 0.168398 | 15 | ns | ns |
| BRIEF-P Plan/Organize | 0.121951 | 30 | 0.288844 | 5 | ns | ns |
| ABC Lethargy | 0.122451 | 42 | 0.217391 | 15 | ns | ns |
| BRIEF-P Behavioral Regulation Index | 0.12786 | 30 | 0.716801 | 5 | ns | ns |
| ABC Stereotypy | 0.179936 | 42 | 0.611838 | 15 | ns | ns |
| BRIEF-P Shift | 0.217766 | 31 | 0.2765 | 5 | ns | ns |
| VABS-2 Daily Living Skills | 0.227381 | 39 | 1 | 12 | ns | ns |
| VABS-2 Adaptive Behavior Composite | 0.271528 | 38 | 0.360765 | 10 | ns | ns |
| ABC Total Score | 0.277263 | 41 | 0.942365 | 15 | ns | ns |
| BRIEF-P Inhibition | 0.553848 | 31 | 0.288844 | 5 | ns | ns |
| VABS-2 Communication | 0.34767 | 39 | 0.931608 | 13 | ns | ns |
| BRIEF-P Monitor | 0.358517 | 29 | 1 | 5 | ns | ns |
| VABS-2 Socialization | 0.37005 | 38 | 0.757315 | 11 | ns | ns |
| ABC Irritability | 0.40207 | 42 | 0.942209 | 15 | ns | ns |
| BVMI Standard Score | 0.467631 | 40 | NA | 2 | ns | ns |
| DCDQ Control During Movement | 0.657687 | 39 | 0.581739 | 15 | ns | ns |
| DCDQ General Coordination | 0.592097 | 39 | 0.706165 | 15 | ns | ns |
| DCDQ Fine Motor | 0.700615 | 39 | 0.738883 | 15 | ns | ns |
| ABC Hyperactivity | 0.758291 | 42 | 0.884722 | 15 | ns | ns |
| VABS-2 Motor Skills | 0.819499 | 18 | 1 | 9 | ns | ns |
| DCDQ Total Score | 0.927195 | 39 | 0.88325 | 15 | ns | ns |

Pairwise domain comparisons of neuropsychological scores. The table presents pairwise Wilcoxon test results comparing PTEN domains (Phosphatase vs C2) and MTOR domains (FAT vs PI3K) for each neuropsychological measure. Sample sizes (n) and significance levels (* $p<0.05$, ** $p<0.01$, ns not significant) are shown for each comparison. **Abbreviations**: VABS, Vineland Adaptive Behavior Scales; SRS, Social Responsiveness Scale; CBCL, Child Behavior Checklist; BRIEF-P, Behavior Rating Inventory of Executive Function - Preschool Version; ABC, Aberrant Behavior Checklist; DCDQ, Developmental Coordination Disorder Questionnaire; BVMI, Beery-Buktenica Developmental Test.

**Figure S1.** Domain-Specific Effects on Additional Neuropsychological Phenotypes

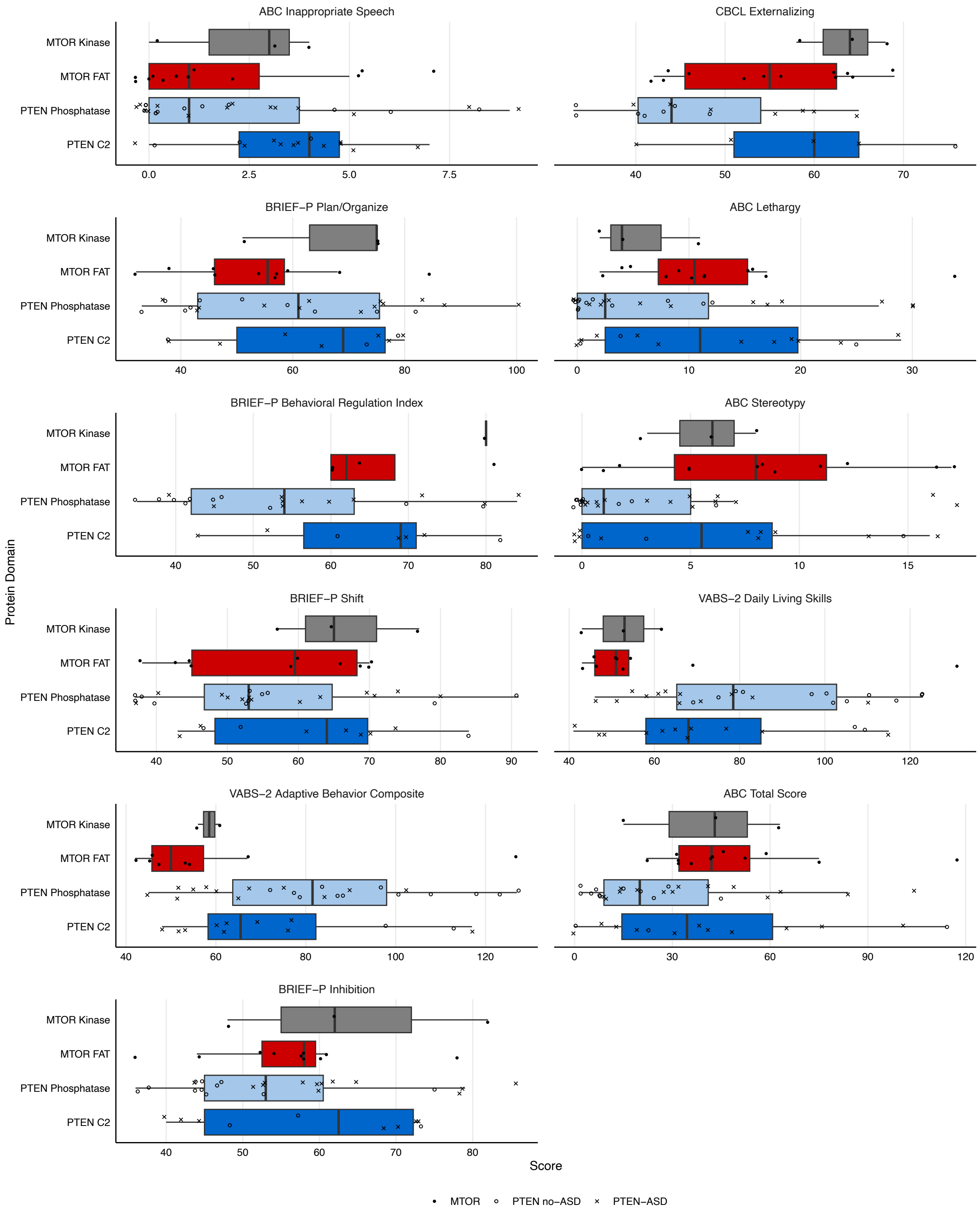

**Figure S1.** Continued

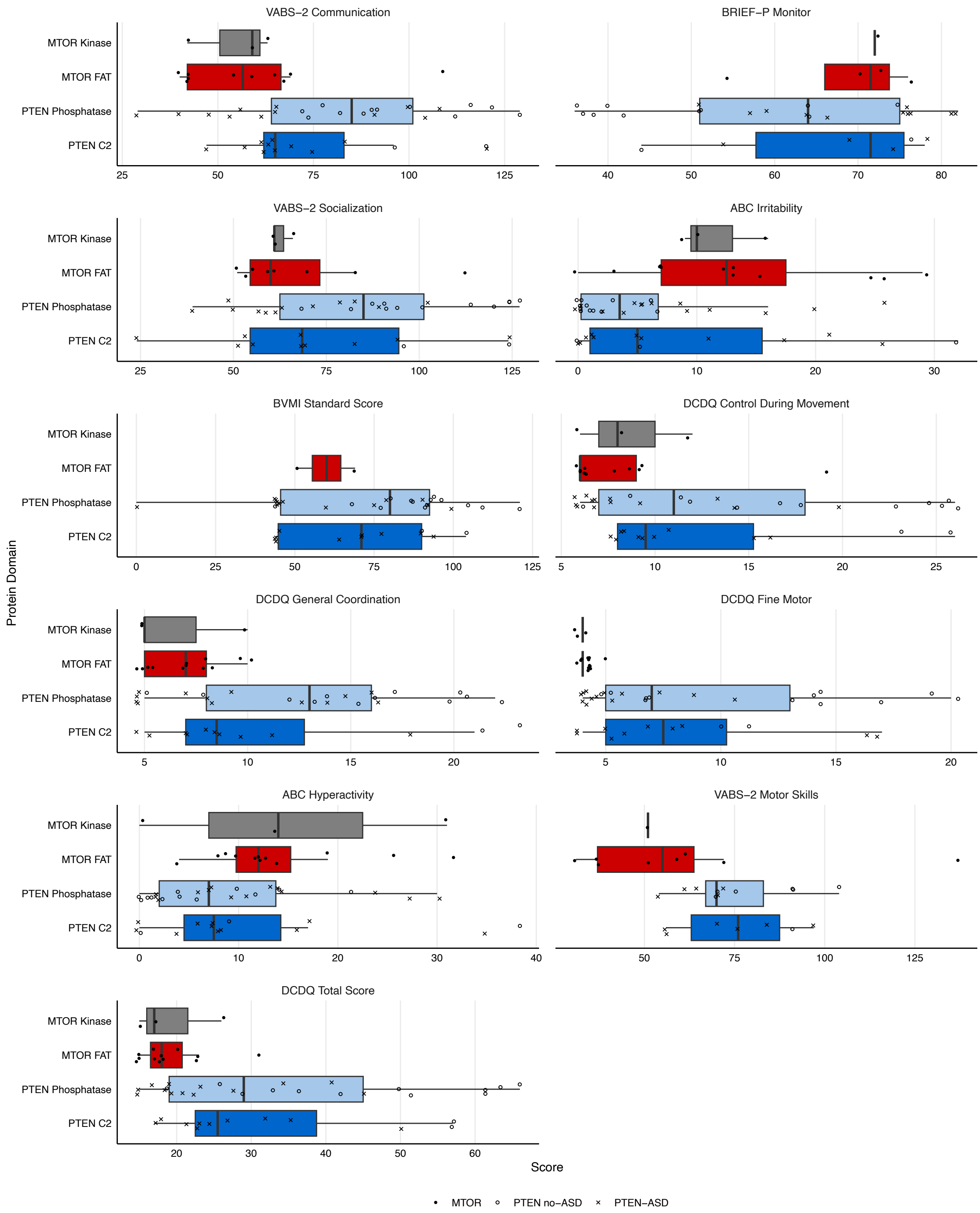


Domain-specific differences in additional neuropsychological phenotypes. Box plots showing the distribution of scores for the remaining twenty-two neuropsychological measures across PTEN and MTOR protein domains. Box plots represent median and interquartile range, with individual data points overlaid. For PTEN variants, filled circles indicate individuals with ASD diagnosis while X's indicate those without ASD. Diamond shapes represent individuals with MTOR variants. Colors indicate protein domains: PTEN Phosphatase (light blue), PTEN C2 (dark blue), MTOR FAT (light red), and MTOR PI3K (dark red). Abbreviations: ABC, Aberrant Behavior Checklist; BVMI, Beery-Buktenica Developmental Test; DCDQ, Developmental Coordination Disorder Questionnaire; VABS, Vineland Adaptive Behavior Scales; BRIEF-P, Behavior Rating Inventory of Executive Function - Preschool Version.
